# Combining Machine Learning with Cox models for identifying risk factors for incident post-menopausal breast cancer in the UK Biobank

**DOI:** 10.1101/2022.06.27.22276932

**Authors:** Xiaonan Liu, Davide Morelli, Thomas J Littlejohns, David A Clifton, Lei Clifton

## Abstract

Breast cancer is the most common cancer in women. A better understanding of risk factors plays a central role in disease prediction and prevention. We aimed to identify potential novel risk factors for breast cancer among post-menopausal women, with pre-specified interest in the role of polygenic risk scores (PRS) for risk prediction.

We designed an analysis pipeline combining both machine learning (ML) and classical statistical models with emphasis on necessary statistical considerations (e.g. collinearity, missing data). Extreme gradient boosting (XGBoost) machine with Shapley (SHAP) feature importance measures were used for risk factor discovery among ∼1.7k features in 104,313 post-menopausal women from the UK Biobank cohort. Cox models were constructed subsequently for in-depth investigation.

Both PRS were significant risk factors when fitted simultaneously in both ML and Cox models (*p* < 0.001). ML analyses identified 11 (excluding the two PRS) novel predictors, among which five were confirmed by the Cox models: plasma urea (HR=0.95, 95% CI 0.92−0.98, *p* < 0.001) and plasma phosphate (HR=0.67, 95% CI 0.52−0.88, *p* = 0.003) were inversely associated with risk of developing post-menopausal breast cancer, whereas basal metabolic rate (HR=1.15, 95% CI 1.08−1.22, *p* < 0.001), red blood cell count (HR=1.20, 95% CI 1.08−1.34, *p* = 0.001), and creatinine in urine (HR=1.05, 95% CI 1.01−1.09, *p* = 0.008) were positively associated.

Our final Cox model demonstrated a slight improvement in risk discrimination when adding novel features to a simpler Cox model containing PRS and the established risk factors (Harrell’s C-index = 0.670 vs 0.665).

## 2. Introduction

Breast cancer is the most common cancer among women, with 2.3 million women diagnosed with breast cancer in 2020 ^1^. Decades of efforts have established multiple risk factors ^2^ for the disease, including reproductive factors ^3–5^, lifestyle ^6,7^, and inherited genetic factors ^8–10^. Despite the identification of multiple modifiable risk factors, breast cancer remains a leading cause of death, with 685,000 deaths in 2020 worldwide. Pre- and post-menopausal breast cancers are usually regarded etiologically different ^11–15^.

Traditionally, risk factor discovery for diseases such as breast cancer is hypothesis-driven. While it is reasonable to use classical statistical models (e.g. logistic regression) to assess these risk factors, some novel risk factors may be overlooked in the discovery stage in information-rich data prior to constructing a classical prediction model. Machine learning (ML) methods are able to handle both a large number of predictors and complex non-linear relationships, hence may provide assistance in the discovery of risk factors ^16,17^. Previous ML studies have primarily focused on how ML approaches compare to conventional models for breast cancer risk prediction cancer ^18–22^, but there are a lack of studies on utilising ML for risk factor identification. The increasing availability of large and detailed cohorts, such as the UK Biobank (UKB), offer the opportunity to utilise hypothesis-free approaches for the identification of potentially novel risk factors.

Recent years have witnessed the rapid development of polygenic risk scores (PRS) which aggregate the effect of a large number (e.g. hundreds or thousands) of genetic variants associated with a specific disease or trait, identified using genome-wide association (GWAS) studies. Breast cancer PRS have been incorporated into existing risk prediction models such as the Breast and Ovarian Analysis of Disease Incidence and Carrier Estimation Algorithm (BOADICEA) ^23^ and Tyrer-Cuzick model ^24^. Interactions between PRS and phenotypic features (e.g. gene-environment interactions) have been suggested with breast cancer and its subtypes, including alcohol consumption, height, hormone therapy ^25^, family history ^26^, hormonal birth control use, menopausal status ^27^, and use of corticosteroids ^28^. However, the overall evidence is inconsistent.

In this paper, we designed an analysis pipeline using two-step approach: ML methods for risk factor discovery, followed by classical Cox models for in-depth investigation ^17,29^. We anticipated differences between our ML and Cox models at the design stage of this study. Our intention was neither to seek superiority among different approaches, nor to build competing prediction models for breast cancer. Our goal was to demonstrate that ML methods can be used to complement classical statistical methods. Furthermore, we used SHapley Additive exPlanation (SHAP) feature dependence plots to explore potential interactions between PRS and phenotypic features. We also provided necessary statistical considerations before constructing classical Cox models to further investigate the novel features discovered by ML methods.

## 3. Methods

### 3.1 Study Design and Participants

The UKB is a large-scale population-based prospective cohort with detailed phenotypic and genetic data from over half a million participants recruited between 2006 and 2010 across 22 assessment centres in England, Wales and Scotland ^30^. The baseline data were collected in person via questionnaires, verbal interview with a trained nurse, physical examinations and biological samples. Follow-up information was obtained through linkage to electronic medical records of death and cancer registries and hospital inpatient records.

In this study, we focused only on post-menopausal women due to the etiological heterogeneity of breast cancer by menopause status ^11–15^. We restricted our study population to a sub-cohort of UKB female participants who were post-menopausal with age 40-69 at baseline, met UKB internal genetic quality control (UKB field 22020), were of genetic White ancestry (UKB field 22006), and had no history of breast cancer, breast carcinoma in situ or mastectomy at baseline (Figure 1).

**Figure 1.**
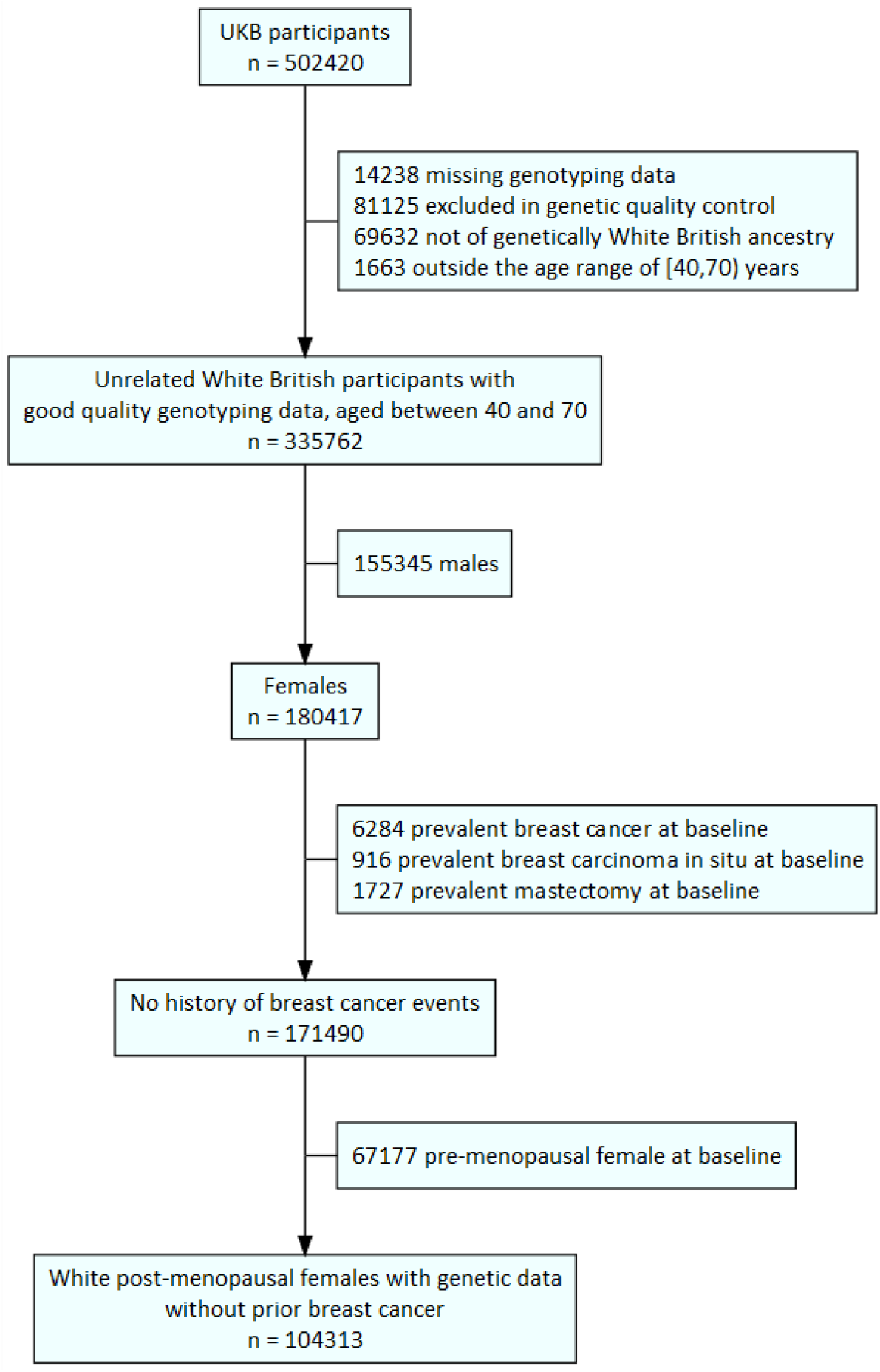
Flowchart illustrating the selection process for our study population.

Our final study population was further randomly divided into training (80%) and test (20%) sets (i.e. training-test split) for subsequent ML analyses.

The UK Biobank study received ethical approval from the North West Multi-center Research Ethics Committee (REC reference: 11/NW/03820). All participants gave written informed consent before enrolment in the study, which was conducted in accordance with the principles of the Declaration of Helsinki. This study has been conducted under the UK Biobank application ID 33952.

### 3.2 Prevalent and incident post-menopausal breast cancer

Prevalent breast cancer cases were identified using International Classification of Diseases codes (ICD-9: 174.X and ICD-10: C50.X) from the linked cancer registry data, with the date of breast cancer diagnosis preceding or on the date of baseline assessment.

Incident cases were identified using cancer registry data, supplemented by record-level hospital inpatient data due to the reporting delay in registries. The follow-up time for each participant was calculated as the number of years from the date of baseline assessment until the earliest of the following: breast cancer diagnosis, date of death from other causes, date of loss to follow-up, date of mastectomy or last date of medical record availability in UKB: 28^th^ February 2021 in England, 28^th^ February 2021 in Scotland, and 28th February 2018 in Wales.

### 3.3 Polygenic risk scores

Due to the substantial discordance in individual-level risk categorisation between different PRS for the same disease ^31^, we included two breast cancer PRS as potential genetic features: PRS_313_^9^ and PRS_120k_^32^. Neither PRS used UKB data in its derivation stage, hence both are suitable for calculation within the UKB population without the concern of inflated effect estimates due to sample overlap. PRS_313_ consisted of 313 (pre-Quality control) independent (correlation < 0.9) genetic variants associated with breast cancer, developed using hard-thresholding and stepwise forward regression with *p* < 10^−5^ in Breast Cancer Association Consortium (BCAC) data. PRS_120k_ consisted of 118,388 (pre-Quality control) variants, developed using the lassosum method ^33^ from the same BCAC data.

We used the imputed genetic data from UKB (version 3, March 2018 release). Full details of genotyping and imputation are described elsewhere ^34^. We performed further variant Quality control (QC) checks across the whole cohort using a published pipeline ^35^, excluding variants that were not available in UKB, variants poorly imputed in UKB (imputation information < 0.4), ambiguous variants (A/T or C/G single nucleotide polymorphisms (SNPs) with minor allele frequency > 0.49) and variants with minor allele frequency (MAF) < 0.005. This led to 305 variants remaining in PRS_313_ and 115,300 in PRS_120k_ (Supplementary Table 1).

We then performed sample QC, excluding participants who were related (third degree or higher), sex discordant, or identified as outliers for genotype missingness or heterozygosity (as these could indicate poor sample quality), using sample QC data provided centrally by UKB (UKB field 22020) that retained a maximal set of unrelated individuals.

Finally, we calculated the PRS as the weighted sum of effect allele dosages, and divided by the number of alleles to account for the vastly different number of variants between the two PRS.

### 3.4 Phenotypic features

In a phenome-wide scan of risk factors for breast cancer, we considered 2,315 features from the UKB (Supplementary Table 2), reflecting socio-demographics, lifestyle, family history, early life and reproductive factors, blood and urine assays, physical measures, cognitive function, medication use and health conditions at baseline.

We mapped the 6,745 unique self-reported medications (UKB field 20003) to 411 distinct codes at level 4 of the Anatomical Therapeutic Chemical (ATC) classification system ^28,36^. For example, if participants self-reported taking “kliofem tablet” or “kliovance 1mg/0.5mg tablet”, they would be categorised into the “G03FA” ATC group (i.e. “contraceptive and hormone replacement therapy (HRT) related medication” group). Since our study population is post-menopausal women only, any G03FA medication is assumed to be HRT, and is referred to as such throughout this paper.

We identified prevalent cancer diagnoses (level 2 ICD-10 codes under “Chapter II Neoplasms”, “Chapter XV Pregnancy, childbirth, and the puerperium”) from cancer registry data, and prevalent non-cancer diagnoses (all ICD-10 codes except Chapters U,V,W,X,Y, and Z) from hospital inpatient data.

We did not include administrative variables, inapplicable pilot fields, male-specific factors, family history of family members of adoptees, history of surgical operations (because they likely reflect existing diagnoses that were already included as input features), and fields collected exclusively during follow-up (e.g. imaging data).

The main pre-processing we performed on training data prior to ML analysis was assigning the following three categories as missing: “Prefer not to answer”, “Do not know”, and empty entries. We then removed features with missing rate > 30%, and those where all participants had the same value (such as rare diseases which no participants were affected by at baseline) which were of no discriminative utility, yielding 1,737 input features for ML models.

### 3.5 Analysis pipeline

We developed an analysis pipeline for combining ML and statistical approaches (Figure 2). The tree-based eXtreme Gradient Boosting (XGBoost) machine learning algorithm ^37^ was used to discover novel features among ∼1.7k variables. Features of high importance according to the SHAP measure were regarded as potentially novel risk factors, and were subsequently investigated by classical Cox models.

**Figure 2.**
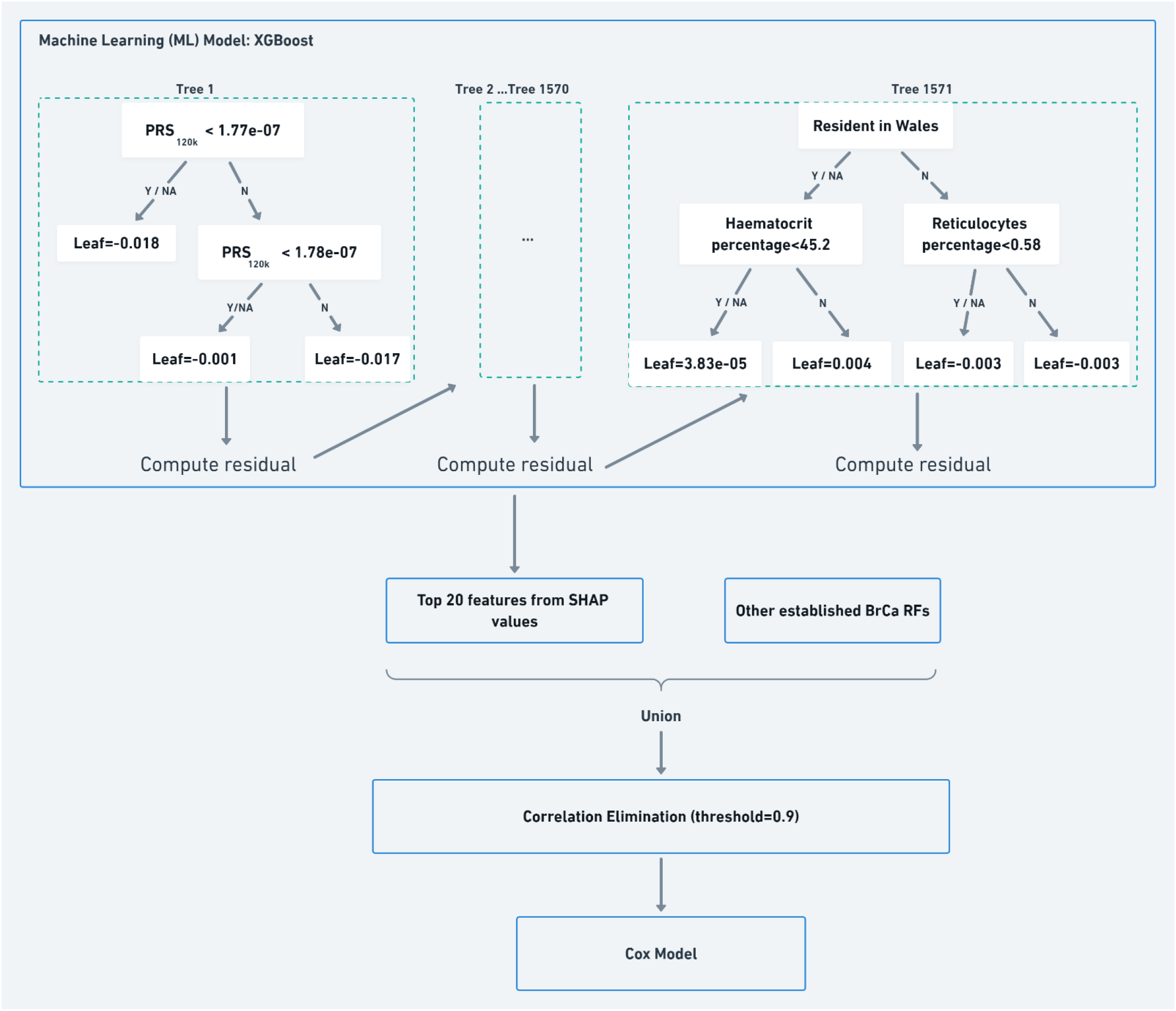
Analysis pipeline. ML models were used for risk factor discovery, followed by classical Cox modelling for further investigation. The example trees derived from our data are shown for illustrative purposes. ML: Machine learning. XGBoost: extreme gradient boosting machines. SHAP: Shapley Additive Explanation. BrCa: Breast cancer. RF: Risk factors. Y/NA: Yes/Missing. N: No.

Our analysis pipeline can be readily generalised to other ML methods in the feature discovery stage. This section describes our analysis pipeline in detail, with an emphasis on the necessary statistical considerations in the process.

### 3.6 ML model

Tree-based XGBoost is an ensemble learning method in which decision trees are built in a sequential manner. After initialising the model prediction by minimising a regularised loss function, the algorithm builds each tree by minimising the residuals from prior ones. The final model prediction is the weighted sum of the predictions from these sequential trees. It is capable of revealing non-linear relationships among correlated features from large datasets in a memory-efficient manner. The advantages of XGBoost over traditional gradient boosting machines (GBM) ^38^ include fast parallel learning, sparsity-aware split finding, and further regularisation that reduces over-fitting and improves model generalisation.

Our outcome (i.e. response variable) was a binary indicator of incident breast cancer (present/absent), with log-loss as the loss function. Missing data was regarded as containing information (i.e. missing not at random (MNAR)). During the training of the model, samples with missing values were assigned a default direction in each branch to either the left or the right child node, based on the gain.

For hyper-parameter tuning, we performed grid search (Supplementary Table 3) with five-fold cross validation (CV) on training data using Area Under the receiver operating characteristic Curve (AUC) as the evaluation metric. The optimal set of hyper-parameters for XGBoost were found to be: maximum depth = 2, number of trees = 1,571, learning rate = 0.01, minimum of child weights = 3, gamma = 0.8, subsample = 0.8, column sample by tree = 0.9, lambda for regularisation = 18, and scale positive weight = 1. For illustrative purpose, the structures of the first and last trees are outlined in Figure 2. The AUC obtained from the five-fold CV was 0.6680 on training data and 0.6679 on test data, indicating that the model was not over-fitted.

A variety of measures exist for obtaining features of high importance, such as the XGBoost built-in methods (“weight” ^37^, “gain” ^39^, “cover”, “total_gain”, and “total_cover”), permutation based feature importance ^40^, and SHAP values ^41^. Existing literature ^17,42^ suggests that SHAP values are the most consistent and stable among the above methods. These properties are vital aspects of feature selection as they provide assurance that the selected features are robust to the perturbation of input data ^43^. SHAP values also have the advantages of faster computation and better visualisation compared to permutation-based methods (Supplementary Materials).

We therefore chose SHAP values as our main feature importance measure, but also implemented the XGBoost default feature importance (“weight”) and permutation-based methods for comparison. The SHAP value of each feature was first computed using one sample at a time to reflect the local effect on the sample, and then aggregated by taking the mean of absolute SHAP values (SHAP_ma_) across all samples to summarise the global attribution of this feature. Global SHAP values of features were presented in a SHAP summary bar plot, whereas local SHAP values were visualised in SHAP dependence plots ^44^ to explore the potential relationship between PRS and phenotypic features.

### 3.7 Statistical model

Following the ML analysis, we further examined the extracted features as follows:

We regarded the top 20 features ranked by SHAP_ma_ as “important”. The union of these 20 features with the established risk factors forms the set of potentially “important” features. We then computed different forms of pairwise correlation *r* among these different types of features from the training data. Spearman’s rank coefficient was computed for pairs of numeric features, and Cramer’s V (computed using the Chi-squared statistic) for pairs of categorical features. The correlation between a numeric and a categorical feature was computed by regressing the numeric feature on the categorical feature and then taking the square root of the proportion of variation explained (also called correlation ratio).

Within each pair of features that was identified as highly correlated (*r* > 0.9), we removed either the feature with most missing data, or the auxiliary one. This step is necessary to reduce the collinearity prior to constructing a linear (e.g. Cox) statistical model.

The missingness within each feature was carefully assessed at this stage, as the number of features had now been sufficiently reduced (e.g. from over 1k to under 30) to permit such close inspection. For example, the variables “Age at first birth” and “Number of live births” needed to be considered together to ensure the imputed data were reasonable for women who have not had children. We performed multiple imputation using the *mice* package in R (Groothuis-Oudshoorn, 2011) to impute the missing data under the assumption of missing at random (MAR). In contrast, the XGBoost machine had assigned a missing category to missing data, effectively assuming MNAR.

We note that the above statistical procedure is essential preparation before constructing classical statistical models and must not be overlooked. The analytical power of their elegant equations comes from careful attention to model specifications and thorough examination of the underlying assumptions.

After the necessary preparation above, we constructed a Cox proportional hazard model to assess the associations between novel features and incident breast cancer, adjusting for established risk factors. Since PRS were present in the model, we also adjusted for genetic array and the first 10 genetic principal components to account for the underlying population structure. As a sensitivity analysis to determine whether the novel features identified by ML improve risk discrimination, we built a separate Cox model without the new features and compared the Harrell’s C-index ^46^ of the two Cox models. Both the fully adjusted and the simpler Cox models were pre-specified in our statistical analysis plan.

The proportional hazards assumption of Cox models was visually assessed using scaled Schoenfeld residuals. Multicollinearity was assessed by computing the variance inflation factor (VIF); values less than 10 were considered acceptable. Statistical tests were two-tailed, and performed using a 5% significance level.

As sensitivity analyses, we built additional Cox models to investigate the potential “PRS × phenotypic features” interactions indicated by the SHAP dependence plots. To further confirm the robustness of feature importance ranking, we implemented another machine learning model, histogram-based gradient boosting machines (GBM) inspired by LightGBM ^47^ (Supplementary Materials).

XGBoost version 1.5.0 and SHAP version 0.40.0 were implemented in Python version 3.8.8 and Cox model analyses were conducted using R version 4.0.2.

## 4. Results

### 4.1 Participants characteristics

Baseline characteristics of the study population are presented in Table 1. Of the 104,313 participants included in our study, 4,010 (3.8%) developed breast cancer over the median follow-up of 11.9 (IQR 11.0-12.6) years. The 80% training and 20% test sets had 3252 and 758 incident cases of breast cancer, respectively.

**Table 1.** Baseline characteristics of the study population by incident breast cancer status (N= 104,313). Median (interquartile ranges, IQR) are presented for continuous variables, frequency (percentage) are reported for categorical variables. Percentages may not add up to 100 due to rounding. N is the number of non-missing values. Note*: Both PRS were multiplied by the number of alleles in the score for easy comparison. BrCa: Breast Cancer. BMI: Body mass index. HRT: hormone replacement therapy. ML: machine learning. MET: Metabolic Equivalent Task. U/L: units per litre.

|  | Without incident BrCa<br>(n=100,303) | With incident BrCa<br>(n=4,010) | N |
| --- | --- | --- | --- |
| *PRS*<sub>120k</sub> * | -0.14 (-0.32, 0.03) | -0.02 (-0.19, 0.16) | 104313 |
| *PRS*<sub>313</sub> * | -0.42 (-0.83, -0.01) | -0.13 (-0.54, 0.27) | 104313 |
| **Established risk factors** |  |  |  |
| Testosterone, nmol/L | 0.97 (0.69, 1.33) | 1.03 (0.75, 1.43) | 81362 |
| Age at recruitment, years | 61.30 (57.08, 64.91) | 61.83 (58.00, 65.14) | 104313 |
| Age at menopause, years | 50.00 (48.00, 53.00) | 51.00 (48.00, 54.00) | 97540 |
| Alcohol units per week | 6.00 (0.00, 12.00) | 6.00 (0.00, 13.50) | 104313 |
| IGF-1, nmol/L | 20.16 (16.56, 23.71) | 20.40 (16.64, 23.89) | 98843 |
| Age at first birth (Categorical) |  |  |  |
| No Births | 16164 (16.1%) | 643 (16.0%) | 16807 |
| <20 | 7616 (7.6%) | 278 (6.9%) | 7894 |
| 20-30 | 60526 (60.4%) | 2371 (59.2%) | 62897 |
| 30-40 | 15181 (15.2%) | 685 (17.1%) | 15866 |
| >=40 | 690 (0.7%) | 30 (0.7%) | 720 |
| BMI, Kg/m<sup>2</sup> | 26.12 (23.54, 29.55) | 26.78 (24.12, 30.32) | 103958 |
| Family history of BrCa | 10807 (10.8%) | 621 (15.5%) | 104313 |
| Summed MET minutes per week | 1786.00 (834.00, 3546.00) | 1667.25 (797.50, 3336.38) | 79981 |
| HRT user | 2257 (2.3%) | 159 (4.0%) | 104313 |
| Age at menarche, years | 13.00 (12.00, 14.00) | 13.00 (12.00, 14.00) | 101470 |
| Number of live births | 2.00 (1.00, 2.00) | 2.00 (1.00, 2.00) | 104261 |
| **Novel predictors selected by ML** |  |  |  |
| Whole body fat mass, Kg | 25.50 (20.20, 32.00) | 26.90 (21.40, 33.70) | 102571 |
| Plasma urea, mmol/L | 5.35 (4.61, 6.18) | 5.31 (4.57, 6.17) | 99337 |
| Basal metabolic rate, KJ | 5489.00 (5138.00, 5912.00) | 5598.00 (5226.00, 6021.00) | 102593 |
| Plasma phosphate, mmol/L | 1.21 (1.12, 1.31) | 1.20 (1.10, 1.29) | 90431 |
| Sodium in urine, mmol/L | 55.20 (35.50, 83.70) | 57.20 (37.73, 86.00) | 100733 |
| Red blood cell count, 10<sup>12</sup>/L | 4.34 (4.12, 4.55) | 4.37 (4.15, 4.58) | 101089 |
| Aspartate aminotransferase, U/L | 23.80 (20.80, 27.60) | 23.60 (20.60, 27.20) | 99091 |
| Creatinine (enzymatic) in urine, mcmol/L | 5530.00 (3412.00, 9027.00) | 5872.00 (3622.50, 9612.00) | 101036 |
| Monocytes count, 10<sup>9</sup>/L | 0.41 (0.33, 0.50) | 0.42 (0.35, 0.52) | 100903 |
| Alkaline phosphatase, U/L | 86.60 (73.10, 102.30) | 86.45 (72.90, 101.60) | 99411 |
| C-reactive protein, mg/L | 1.42 (0.69, 2.95) | 1.61 (0.79, 3.29) | 99211 |

### 4.2 Input features for ML

Figure 3 shows the categories of the 1,737 input features for the XGBoost ML models, over half of which were in the “Health conditions” category (e.g. infectious diseases, circulatory diseases, and cancers). “Lifestyle factors” include alcohol, diet, and sleep. “Medication use” includes medication for blood pressure control, and birth control. “Physical measures” include blood pressure, arterial stiffness, and anthropometry. “Socio-demographics” include age, education, employment, and deprivation index. “Blood and urine assays” include blood counts and biochemistry (e.g. cholesterol). “Early life and reproductive factors” include birth weight and age at menarche. “Family history” includes illnesses of father, mother, and siblings.

**Figure 3.**
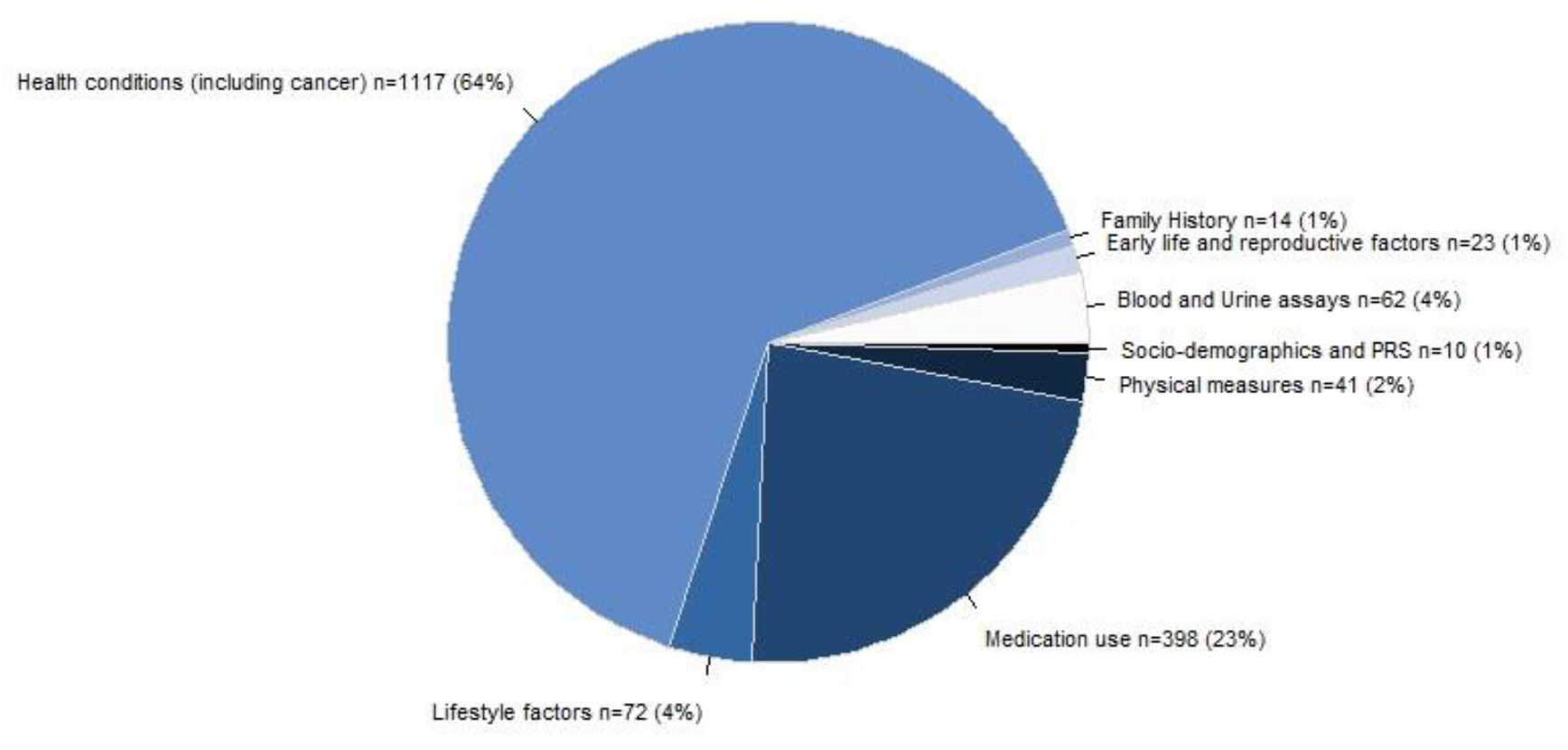
Categories of the 1,737 input features for ML analyses; n (%) represents the number (percent) of features included in each category

### 4.3 ML analysis

The top 20 features ranked by the highest mean absolute SHAP values (SHAP_ma_) are shown in Figure 4 (full list of SHAP_ma_ in Supplementary Excel file). There are a total of 14 established risk factors available in the UKB. Two of these are PRS_120k_ and PRS_313_, occupying the top two positons, with noticeably higher feature importance values than the remaining features, indicating that both PRS warrant inclusion in the subsequent models. Seven of these (testosterone, age, age at menopause, family history of breast cancer, alcohol intake, IGF-1, age at first birth) appeared in the top 20 features. Five of these not shown in Figure 4 are physical activity (i.e. Summed MET minutes per week) (ranked 23, SHAP_ma_ = 0.016), use of HRT (ranked 25, SHAP_ma_ = 0.015), body mass index (BMI) (ranked 66, SHAP_ma_ = 0.004), age at menarche (ranked 76, SHAP_ma_ = 0.003), and parity (ranked 88, SHAP_ma_ = 0.002).

**Figure 4.**
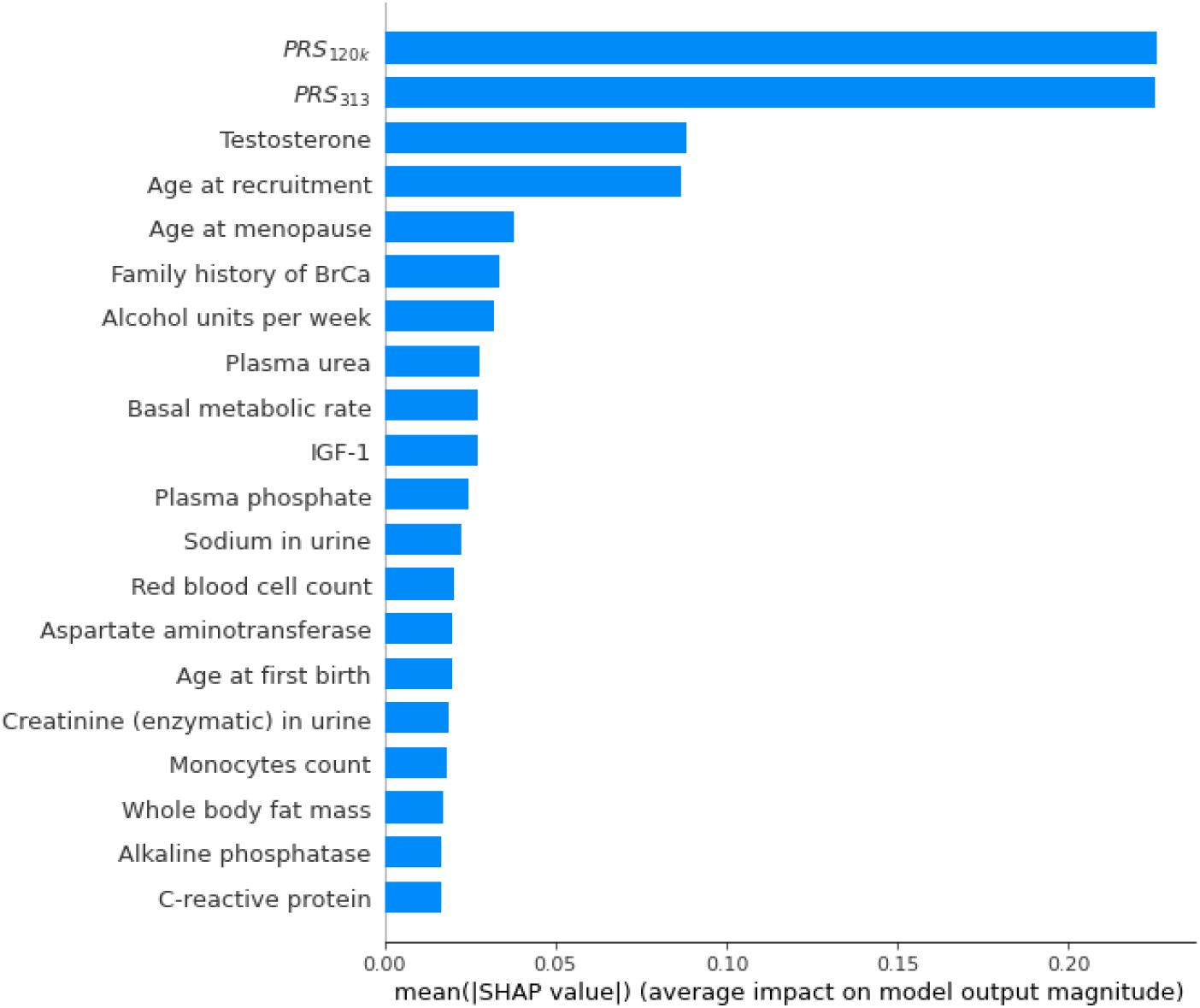
SHAP summary bar plot showing the top 20 most important features for the risk of breast cancer, according to the XGBoost machine and SHAP values. Noticeably, both BrCa PRS are deemed of much higher importance than the remaining phenotypic features. BrCa: Breast Cancer. SHAP: SHapley Additive explanation. mean(|SHAP value|): mean absolute SHAP value, SHAP_ma_.

Our XGBoost machine discovered novel predictors of breast cancer from the following categories among its top 20 ranked features:

- Body composition by impedance (UKB Category 100009) (basal metabolic rate, whole body fat mass)
- Blood count (UKB Category 100081) (red blood cell count, monocytes count)
- Blood biochemistry (UKB Category 17518) (plasma urea, plasma phosphate, aspartate aminotransferase, alkaline phosphatase, C-reactive protein)
- Urine assays (UKB Category 100083) (sodium in urine, creatinine in urine)

BMI was highly correlated with whole body fat mass (*r* = 0.92). Given that both variables had similar missingness, we removed BMI and kept whole body fat mass in further investigations, because whole body fat mass was regarded as a more accurate measure for capturing body composition, and was ranked higher than BMI by the SHAP value. Another anthropometric measure, basal metabolic rate (measured using a Tanita BC418MA body composition analyser), showed expected correlation with BMI (*r* = 0.73) and whole body fat mass (*r* = 0.76). We retained basal metabolic rate in the subsequent Cox models, because these correlations are below the threshold of 0.9.

During the training of our XGBoost machine, hyper-parameter tuning indicated that allowing each tree to grow down to two levels yielded the best model performance, which in turn allowed the discovery of two-way interactions among features. Given our particular interest in PRS, we used SHAP dependence plots to visualise the main effect of each of the top 20 features and the effect modification by PRS (i.e. interaction with PRS) (Supplementary Fig. 3-4). The SHAP dependence plots revealed potential effect modifications but with some unexpected patterns, which required further investigations before drawing inference on effect modifications. Full results and investigation are presented in Supplementary Materials.

### 4.4 Statistical analysis

Our final multivariable Cox model consists of the 14 established risk factors available in the UKB (including the two PRS), the 11 potentially novel features identified by SHAP value rankings, genetic array, and the first 10 PCs. Among these 11 novel features, the following five had a statistically significant association with breast cancer in post-menopausal women: basal metabolic rate, red blood cell count, plasma urea, plasma phosphate, and creatinine in urine. Among these five features, blood biochemistry features (i.e. plasma urea and plasma phosphate) were inversely associated with risk of developing breast cancer, whereas other novel features (i.e. basal metabolic rate, red blood cell count, and creatinine in urine) were positively associated.

The remaining six novel features that did not reach the 5% significant level are: sodium in urine, aspartate aminotransferase, monocytes count, whole body fat mass, alkaline phosphatase, and C-reactive protein. Figure 5 shows the hazard ratios (HR) with 95% confidence interval and p-values of covariates in the final Cox model.

**Figure 5.**
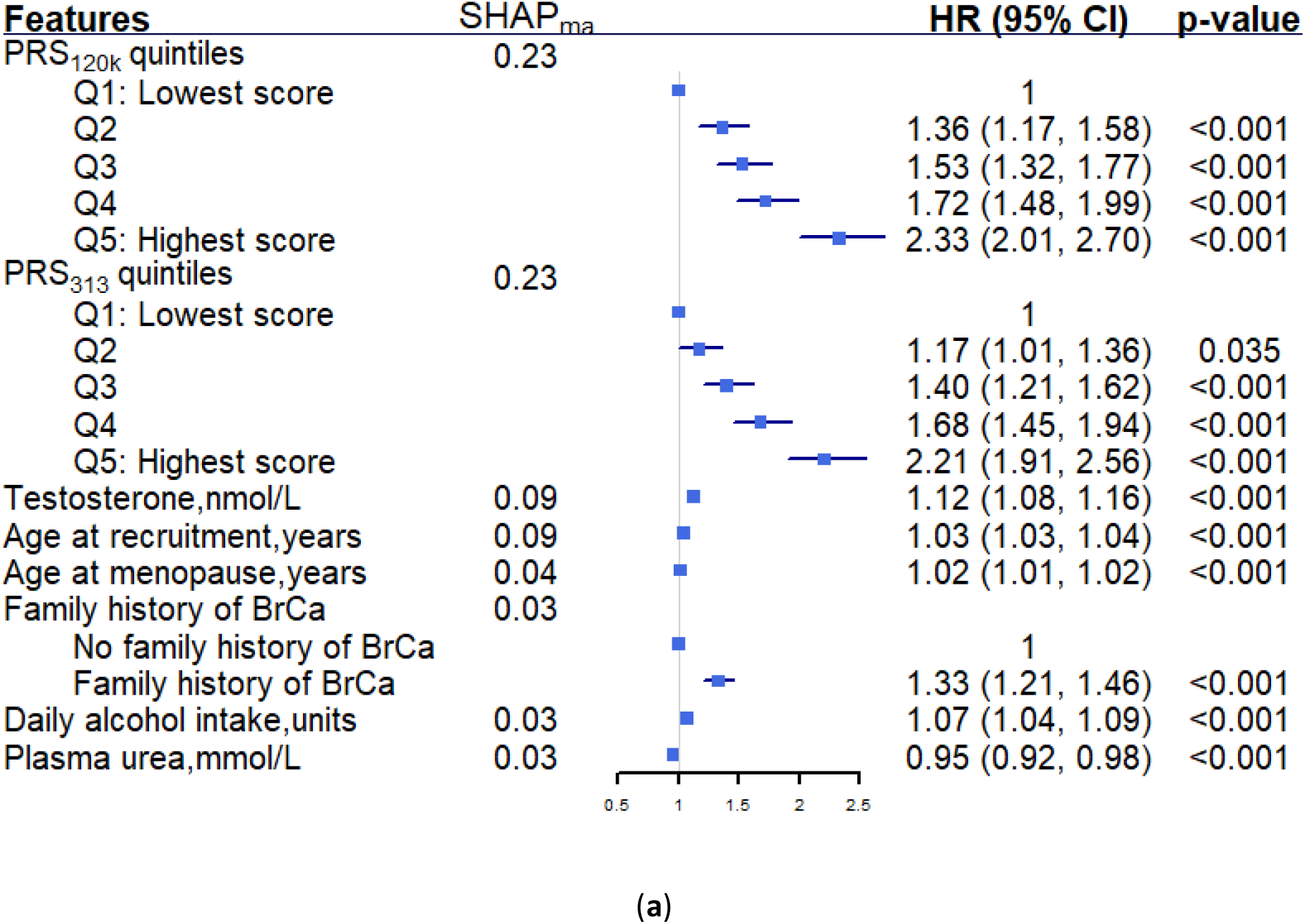
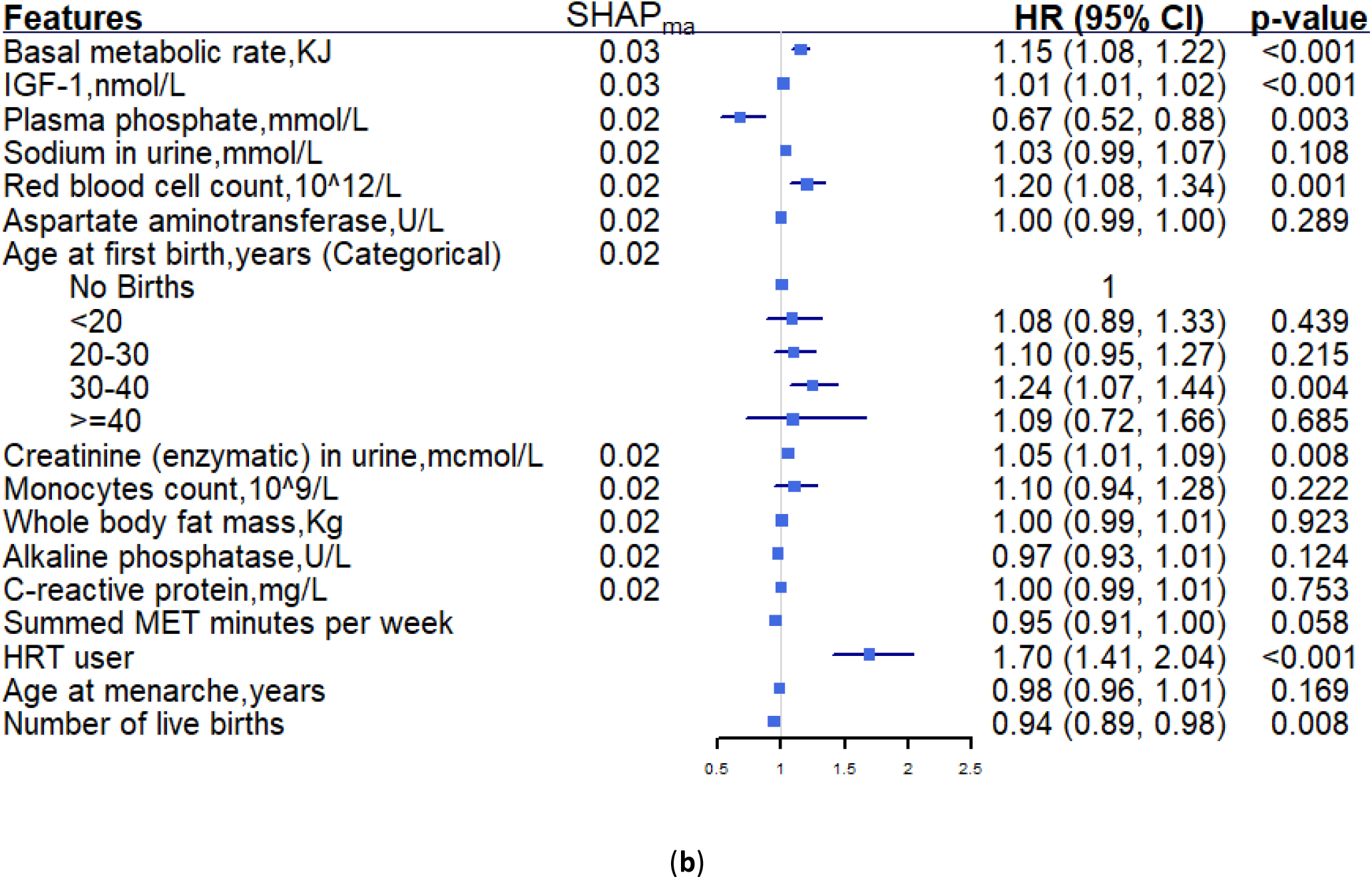
Results obtained from our final multivariable Cox model, separated into two subplots for ease of reading: (a) top 8 features ranked by SHAP_ma_, (b) the remaining 16 features, of which the bottom four features are established risk factors that are outside the top 20 features by SHAP_ma_. Both PRS were categorised into quintiles, alcohol intake was scaled from weekly intake to daily intake for easy interpretation and direct comparison with existing literature. Basal metabolic rate, sodium in urine, creatinine in urine, alkaline phosphatase, and summed MET minutes per week were standardised using the mean and standard deviation within each imputed dataset, hence the corresponding HR represents per 1 standard deviation increase. For other continuous variables, HR represents per 1 unit increase. Genetic array and first 10 PCs were adjusted in the model but omitted from the figure. SHAP: SHapley Additive explanation. SHAP_ma_: mean absolute SHAP value. BrCa: Breast Cancer. HR: hazard ratio. CI: confidence interval. HRT: hormone replacement therapy. MET: Metabolic Equivalent Task. U/L: units per litre.

As a pre-specified sensitivity analysis, we constructed a separate simpler Cox model containing only PRS and the 12 established risk factors (Supplementary Table 5). When adding the additional novel features selected by ML, Harrell’s C-index increased from 0.665 to 0.670 in training data, and from 0.660 to 0.661 in test data.

To interrogate the possibility of reverse causation, we conducted another sensitivity analysis to exclude the first two years of follow-up for blood and urine biomarkers (*N* = 82,491). We did not find evidence of reverse causation (Supplementary Fig. 2).

## 5. Discussion

Our XGBoost machine searched across ∼1.7k features in UKB, and discovered 11 potentially novel risk factors, as well as established ones (e.g. PRS, age, age at first birth, family history of breast cancer, testosterone, and IGF-1). These 11 novel factors came from diverse categories, including body composition measure, blood count, blood biochemistry, and urine assays. Five (basal metabolic rate, red blood cell count, plasma urea, plasma phosphate, and creatinine in urine) of these were confirmed to be statistically significant by the subsequent Cox model.

### 5.1 Mechanism behind the novel risk factors

Here we delineate the potential mechanisms behind the novel factors. Body composition measures were expected to be associated with breast cancer as obesity is a well-known risk factor for post-menopausal breast cancer ^48^. The unexpected observation was that the XGBoost model selected a number of more detailed body composition measures (e.g. basal metabolic rate, whole body fat mass) instead of BMI, indicating that more precise body composition measures could provide important information above and beyond BMI for predicting breast cancer. Previous studies ^49–51^ have investigated anthropometric factors beyond BMI (e.g. waist to hip ratio, weight gain, waist circumference) and found positive associations with post-menopausal breast cancer. However more detailed anthropometric factors are worthy of further investigation. We were not able to confirm our hypothesis in classical Cox models due to collinearity among these anthropometric measures, but future research could deploy different models to dissect this hypothesis. Our findings showed that basal metabolic rate is a significant predictor for breast cancer, contradicting previous studies that found no such association ^52,53^. Our positive finding could be due to the statistical power conferred by the large sample size of UKB.

The SHAP feature importance ranking supported the associations of novel biomarkers with post-menopausal breast cancer, but little literature exists on this topic. While absolute monocytes count was identified as a potential prognostic factor for breast cancer ^54^, the association of red blood cell count with breast cancer has not been studied explicitly. Plasma urea, a blood biomarker related to kidney function, was reported to have null causal relationship with breast cancer ^55^, but our study suggests it may be associated with breast cancer. Aspartate aminotransferase and alkaline phosphatase are both blood biomarkers related to liver function and were not previously associated with non-metastatic breast cancer ^56^. In contrast, C-reactive protein (CRP), a marker of systemic inflammation, was reported to be associated with breast cancer via meta-analysis ^57,58^. To our knowledge, no previous studies have reported the association of plasma phosphate, sodium in urine or creatinine in urine with breast cancer.

Our findings of novel risk factors should be treated with caution, and further examined in independent datasets. These novel features could be surrogates for other processes that are not modelled in our analyses. There is still the possibility of chance findings by the ML model, although we have pre-specified our classical Cox models. We did not observe evidence of reverse causation, but such absence of evidence should not be regarded as evidence of absence.

### 5.2 Undiscovered well-established risk factors

Five well-established risk factors are unavailable (mammographic density, plasma oestrogen, progesterone) or unusable (plasma oestradiol) in the UKB. Plasma oestradiol is measured in UKB, but the measured concentrations of nearly all post-menopausal women were below the reportable range ^13^, hence were regarded as missing values and could not be included in the analysis.

We identified 19 established risk factors for post-menopausal breast cancer from the literature, 14 of which are available in UKB. Five out of these 14 risk factors were not ranked in the top 20 by the SHAP values: BMI was ranked at 66, probably because its related factor (e.g. whole body fat mass) were already ranked among the top 20; Physical activity and HRT use were on the borderline of inclusion in top 20 (ranked 23 and 25 respectively), whereas age at menarche and parity were ranked further behind (ranked 76 and 88 respectively).

This highlights the need for establishing a criterion to decide which features should be regarded as important. Some suggested considering 3% of total number of features as important ^59^, while others advocated a cut-off value of 0.05 for the SHAP feature importance measure ^17^. We pre-specified the top 20 features due to the practical need for keeping the number of features in a manageable range, in the absence of established criteria on this empirical choice.

### 5.3 PRS

We had an a priori interest in PRS, and intended to explore the relationship between the two PRS for breast cancer based on the existing understanding of their correlation. We discovered that both PRS were ranked as the strongest risk factors by the agnostic ML methods, which is surprising given that both PRS were developed using largely the same GWAS data for the same disease. We then conducted in-depth Cox regression simultaneously fitting both PRS as predictors, and concluded that both PRS are significant predictors for post-menopausal breast cancer. This raises the general question of whether multiple PRS should be used to improve risk prediction obtained from a single PRS.

SHAP dependence plots are useful for (i) visualising non-linear relationships between features and outcome, and (ii) revealing potential pairwise interactions between features. However, in this study, we noticed unexpected patterns arising from these plots, indicating that careful investigations are required before drawing firm conclusions.

### 5.4 From ML to classical statistical models

ML methods are well suited for large-scale feature selection, and ongoing methodological development is making ML models more interpretable. It is beneficial to incorporate classical statistical models in feature selection for the following reasons. First, compared to feature importance ranking, statistical models (e.g. Cox regression) are able to quantify the strength of association of each feature with the outcome (e.g. via hazard ratio with 95% confidence interval) with straightforward interpretation. Second, model coefficients (i.e. betas) and p-values are typically used to infer feature importance in classical models, providing a different perspective from the feature importance measure in the ML setting (e.g. SHAP values utilise the impact in model predictions with and without a particular feature). Third, feature importance measures by ML methods may select spurious features purely due to confounding ^17^, and therefore should be further examined in classical statistical models.

When the search scope includes highly correlated features, one needs to carefully choose ML (e.g. tree-based) models that are capable of handling correlations, and then perform correlation and collinearity checks before including the selected features in classical statistical models.

We did not expect the results obtained from an ML method to be fully compatible with those from a classical statistical model, because each approach has its own strengths and limitations. Instead, we aimed to complement Cox models with the insight from the XGBoost machine in this study. There are several possible explanations on the observed differences between the ML method and classical Cox models in this study.

First, our XGBoost machine characteristically makes binary splits of the input features among thousands of trees (Figure 2) assuming non-linear relationship among features, whereas our Cox models are essentially linear. Although it is possible to incorporate non-linearity in Cox models (e.g. using splines or fractional polynomials), it would result in an overly complex model for interpretation. It is neither necessary nor appropriate to anticipate full agreement between ML and classical statistical models. Second, the criteria for inferring feature importance are different between ML and classical models, as described above.

Finally, we emphasise that while agreement between ML and classical models raises the confidence of discovery, differences do not necessarily imply superiority of either approach or compelling us to choose one over the other. The differences could serve as a signal for further investigations where critical thinking should be exercised.

### 5.5 Challenges and solutions

It is worth highlighting the challenges we encountered when implementing our analysis pipeline, and providing potential solutions here. The main challenge is how to handle missing data when combining ML and classical statistics for interpretation. Existing literature ^60,61^ has compared various imputation strategies (e.g. response augmentation framework, K-nearest neighbour, mean imputation) and suggested utilising the method that yields the best prediction accuracy depending on the dataset. However, the potential biases arising from missing data must not be overlooked, particularly in the context of statistical modelling.

It is difficult to keep a consistent approach for handling missing data between ML and classical statistics. In classical statistics, one usually performs multiple imputation that generates multiple completed datasets (usually around 10-20), fits the model using each dataset, and then pools model estimates using Rubin’s rule ^62^. However, it is not feasible to train a ML model on each imputed dataset due to the extensive computation time this would require. One alternative is to perform a single imputation, but this was not compatible with our complex variables where some are missing at random and some are not. For example, “Age at first birth” is not missing at random and if one blindly performs a single imputation, the imputed data would not make sense for women who have not had children. Although manual inspections can be performed on small datasets, it is practically impossible to carefully assess the missing mechanisms of the high (∼1.7k) dimensional data in our study.

Our solution is as follows: For the ML analysis, we treated missing data as a separate category and used the default setting for XGBoost (i.e. when the value needed for the split was missing, a default direction was assigned with the maximum gain). Once we had reduced the number of features to a workable size (∼20), we carefully inspected the missingness and performed multiple imputation as appropriate in the classical statistical setting. One might argue that the missing mechanism for each feature should be consistent throughout the analyses, but such purity is impractical when dealing with large datasets. This issue could be a potential topic for future research.

### 5.6 Strengths and limitations

We performed an agnostic search of potential risk factors for breast cancer in post-menopausal women among ∼1.7k features. We developed an analysis pipeline for combined ML and classical statistical models, and incorporated necessary statistical considerations in our pipeline while acknowledging the anticipated inconsistency between different models. The presence of well-established risk factors, the large sample size, and the long follow-up period of UKB data have enabled us to perform rigorous analyses in the process.

Our study has several limitations. A few well-known risk factors (e.g. mammographic density, plasma oestrogen, progesterone, plasma oestradiol) and detailed family history data were either unavailable or unusable in UKB, hence could not be investigated in our study. We did not investigate subtypes (Estrogen-receptor [ER]-positive or negative) of breast cancer, due to incomplete data on tumour type in UKB. We did not incorporate exome data that are necessary for identifying BRCA1/BRCA2 carriers, or other high penetrance variants. Our study population consists only of genetically white individuals, and therefore should be not generalised to other ethnicities without further research. Finally, we did not compare our model with existing risk prediction models for breast cancer, such as BOADICEA ^23^, Cuzick model ^24^, due to their complexity and incomplete mapping to UKB variables.

## 6. Conclusion

In conclusion, combining ML with Cox models, we identified five statistically significant novel association with post-menopausal breast cancer for blood counts, blood biochemistry and urine biomarkers. We demonstrated a slight improvement in risk discrimination (Harrell’s C-index) when adding these five novel features to the Cox model that only contains PRS and established risk factors. We discovered that both of our pre-specified PRS were ranked as the most important features by SHAP value, and can be simultaneously included in our final Cox model. Our findings support further investigation on using more precise anthropometry measures for improved breast cancer prediction. Other directions for future research include the utility of multiple PRS for better risk prediction, and validating the link between blood/urine biomarkers and risk of breast cancer.

## 7. Data availability

The data reported in this paper are available via application directly to the UK Biobank, *https://www.ukbiobank.ac.uk*.

The code used for analyses are available at *https://github.com/xiaonanl1996/MLforBrCa*.

## Data Availability

All data produced in this paper are available via application directly to the UK Biobank, https://www.ukbiobank.ac.uk. The code used for analyses are available at https://github.com/xiaonanl1996/MLforBrCa.

## 9. Acknowledgements

We thank the participants of the UK Biobank study for enabling us to conduct this research. Computing of this study used the Oxford Biomedical Research Computing (BMRC) facility, a joint development between the Wellcome Centre for Human Genetics and the Big Data Institute supported by Health Data Research UK and the NIHR Oxford Biomedical Research Centre. The views expressed are those of the author(s) and not necessarily those of the NHS, the NIHR or the Department of Health.

## 11. Author contributions

LC conceived the research idea and outlined the methods; XL and LC drafted the manuscript; XL conducted the analyses; TJL reviewed the manuscript; DM and DAC provided consultation on ML methods. All authors have revised the manuscript and agreed on its contents.

## 12. Additional Information

### 12.2 Patient and community involvement

The analyses presented here are based on existing data from the UK Biobank cohort study, and the authors were not involved in participant recruitment. To the best of our knowledge, no patients were explicitly engaged in the design or implementation of the UK Biobank study. No patients were asked to advise on interpretation or writing these results. Results from UK Biobank are routinely disseminated to study participants via the study website and social media outlets.

### 12.3 Consent for publication

Yes.

### 12.4 Transparency statement

The lead author affirms that this manuscript is an honest, accurate, and transparent account of the study being reported; that no important aspects of the study have been omitted; and that any discrepancies from the study as planned have been explained.

