## Supplementary Materials for "Combining Machine Learning with Cox models for identifying risk factors for incident post-menopausal breast cancer in the UK Biobank"

Xiaonan Liu^1*^, Davide Morelli^2^, Thomas J Littlejohns^1^, David A Clifton^2^, Lei Clifton^1^

Affiliate Institutions:

1. Nuffield Department of Population Health, University of Oxford, Oxford, UK.

2. Department of Engineering Science, University of Oxford, Oxford, UK.

### 1. Genetic QC

Supplementary Table 1. Number of SNPs excluded during SNP quality control (QC) of PRS for breast cancer. PRS: Polygenic risk scores. nSNPs: Number of single nucleotide polymorphisms (SNPs) included in the PRS prior to SNP QC. UKB: UK Biobank.

| PRS | nSNPs | Unavailable in UKB | Ambiguous | Imputation info < 0.4 | MAF < 0.005 | Remaining SNPs |
| --- | --- | --- | --- | --- | --- | --- |
| PRS_313_ | 313 | 7 | 0 | 0 | 1 | 305 |
| PRS_120k_ | 118,388 | 0 | 43 | 107 | 2938 | 115,300 |

### 2. Input features

Supplementary Table 2. Input features to the machine learning (ML) model for risk factor discovery of post-menopausal breast cancer (BrCa). The “Category” column specifies variables at the "level 2" category defined by UK Biobank (UKB) (see [UKB website](https://biobank.ndph.ox.ac.uk/showcase/browse.cgi)). The “Sub-category” column specifies variables at the "level 3" category by UKB. The “Variables” column lists the variables selected for consideration with justification in the “Comments” column. FID: Field ID in UKB. Symbol "--" stands for "not applicable" or "not included". Pre-processing on missing categories (i.e. “Prefer not to answer”, “Do not know” and empty entry) were applied to all variables below.

| Category | Sub-category | Variables | Comments |
| --- | --- | --- | --- |
| PRS | -- | PRS_313_ and PRS_120k_ | We computed PRS using imputed genetic data from UKB (version 3, March 2018 release) via pipeline ^1^. Full details in the manuscript. |
| [Socio-demographics category](https://biobank.ndph.ox.ac.uk/showcase/label.cgi?id=701) | [Baseline characteristics](https://biobank.ndph.ox.ac.uk/showcase/label.cgi?id=100094) | Age at baseline, Townsend deprivation score | We did not include gender and ethnicity in the input features, because our study population contains only genetically White females.  We combined FID 6138 and 10722 into one variable called “Highest qualifications achieved”. |
|  | [Education](https://biobank.ndph.ox.ac.uk/showcase/label.cgi?id=100063) | Qualifications |  |
|  | [Employment](https://biobank.ndph.ox.ac.uk/showcase/label.cgi?id=100073) | Employment status, [employment category](https://biobank.ndph.ox.ac.uk/showcase/field.cgi?id=20277) |  |
|  | [Household](https://biobank.ndph.ox.ac.uk/showcase/label.cgi?id=100066) | Household (pre-tax) income |  |
|  | [Indices of Multiple deprivation](https://biobank.ndph.ox.ac.uk/showcase/label.cgi?id=76) | -- |  |
| [Lifestyle category](https://biobank.ndph.ox.ac.uk/showcase/label.cgi?id=704) | [Alcohol](https://biobank.ndph.ox.ac.uk/showcase/label.cgi?id=100051) | Alcohol drinking status, weekly alcohol consumed units | We derived weekly alcohol consumed units by combining consumption from red wine, white wine, beer, cider, spirits, fortified wine and other alcohol. |
|  | [Diet](https://biobank.ndph.ox.ac.uk/showcase/label.cgi?id=100052) | Variables within whole category | Some variables (e.g. [FID 1309](https://biobank.ndph.ox.ac.uk/showcase/field.cgi?id=1309): Fresh fruit intake) contain special values (e.g. “-10” represents “Less than one”), which were coded as 0. |
|  | [Electronic device use](https://biobank.ndph.ox.ac.uk/showcase/label.cgi?id=100053) | Weekly usage of mobile phone in last 3 months | We excluded pilot field [FID 10749](https://biobank.ndph.ox.ac.uk/showcase/field.cgi?id=10749) because its categories do not match with main field (FID 1120). |
|  | [Physical activity](https://biobank.ndph.ox.ac.uk/showcase/label.cgi?id=100054) | Variables within whole category | All pilot fields were omitted because their categories are different from those of the main fields, which made them impossible to be combined. |
|  | [Sleep](https://biobank.ndph.ox.ac.uk/showcase/label.cgi?id=100057) | Variables within whole category | These include sleep duration, getting up in morning, morning/evening person, napping during day, insomnia, snoring, daytime dozing/sleeping. |
|  | [Smoking](https://biobank.ndph.ox.ac.uk/showcase/label.cgi?id=100058) | Smoking status | We used raw UKB variable. |
|  | [Social support](https://biobank.ndph.ox.ac.uk/showcase/label.cgi?id=100061) | Leisure/social activities  Able to confide, frequency of friend/family visits | We used raw UKB variables. |
|  | [Mental health](https://biobank.ndph.ox.ac.uk/showcase/label.cgi?id=100060) | -- | We did not use this self-report category, and instead used ICD-10 codes from “Chapter V Mental and behavioural disorders”. |
|  | [Sun exposure](https://biobank.ndph.ox.ac.uk/showcase/label.cgi?id=100055) | Variables within whole category | We used raw UKB variables. |
| [Family History](https://biobank.ndph.ox.ac.uk/showcase/label.cgi?id=705) | -- | Family history (FaH) of CVD (high BP, stroke, heart disease), FaH of breast cancer, FaH of severe depression, FaH of diabetes, FaH of other cancer (bowel cancer, prostate cancer, lung cancer), FaH of dementia, FaH of Parkinson’s disease, FaH of chronic bronchitis  Variables within rest of the whole category (except fields related to adopted family members) | FaH variables were derived from [illness of father](https://biobank.ndph.ox.ac.uk/showcase/field.cgi?id=20107), [mother](https://biobank.ndph.ox.ac.uk/showcase/field.cgi?id=20110), [siblings](https://biobank.ndph.ox.ac.uk/showcase/field.cgi?id=20111).  We did not incorporate variables regarding adopted father, mother and siblings. |
| [Early life and reproductive factors](https://biobank.ndph.ox.ac.uk/showcase/label.cgi?id=708) | -- | Variables within [Female-specific factors](https://biobank.ndph.ox.ac.uk/showcase/label.cgi?id=100069) and [Early Life factors](https://biobank.ndph.ox.ac.uk/showcase/label.cgi?id=100033) | Male-specific factors were not considered.  Female-specific variables that contain special values were coded as follows:  Value "-10" in variable FID 2704: “[Years since last cervical smear test](https://biobank.ndph.ox.ac.uk/showcase/field.cgi?id=2704)” represents less than a year ago, which was treated as 0.  Value "-6" in variable FID 3710: “[Length of menstrual cycle](https://biobank.ndph.ox.ac.uk/showcase/field.cgi?id=3710)” represents irregular cycle, which was treated as missing.  Value "-4" in variable FID 2764: “[Age at last live birth](https://biobank.ndph.ox.ac.uk/showcase/field.cgi?id=2764)” represents "Do not remember", which was treated as missing. |
| [Health conditions](https://biobank.ndph.ox.ac.uk/showcase/label.cgi?id=100091) | -- | Indicators of each [level 2 of Diagnoses ICD 10](https://biobank.ndph.ox.ac.uk/showcase/field.cgi?id=41270) (except unspecific categories (“Chapter XX”, “Chapter XXI”, “Chapter XXII”) | When deriving health conditions in UKB, one usually investigates multiple resources (e.g. ICD-10, ICD-9, self-report), and then defines the condition across them. However, in this study, we do not have pre-specified health conditions, and it is impractical to combine multiple resources for the definition of each of the 19k conditions present in UKB. Therefore, we utilised one dominant field, [hospital inpatient ICD-10 code](https://biobank.ndph.ox.ac.uk/showcase/field.cgi?id=41270), for medical condition diagnoses at baseline. |
| [Genotype results, process and QC](https://biobank.ndph.ox.ac.uk/showcase/label.cgi?id=716) | -- | -- | We used this category for computing PRS but fields within this category were not considered by the ML model. |
| [Procedural metrics](https://biobank.ndph.ox.ac.uk/showcase/label.cgi?id=718) | -- | -- | This category consists of admin variables, hence not included as input features. |
| [Medication](https://biobank.ndph.ox.ac.uk/showcase/field.cgi?id=20003) | -- | Self-report medication use within each ATC group at baseline from [Treatment/medication code](https://biobank.ndph.ox.ac.uk/showcase/field.cgi?id=20003) | Full details in the manuscript. |
| [Blood Assays](https://biobank.ndph.ox.ac.uk/showcase/label.cgi?id=100080) |  | [Blood biochemistry](https://biobank.ndph.ox.ac.uk/showcase/label.cgi?id=17518), [Blood counts](https://biobank.ndph.ox.ac.uk/showcase/label.cgi?id=100081) | Variable “[Rheumatoid factor](https://biobank.ndph.ox.ac.uk/showcase/field.cgi?id=30820)”, variable “[Oestradiol](https://biobank.ndph.ox.ac.uk/showcase/field.cgi?id=30800)”, all variables in category “[Infectious disease](https://biobank.ndph.ox.ac.uk/showcase/label.cgi?id=51428)”, all variables in category “[Metabolics](https://biobank.ndph.ox.ac.uk/showcase/label.cgi?id=220)” were not considered due to extremely high missing data.  (“Rheumatoid factor” was available for 8% of participants, “Oestradiol” was available for 15% of participants, and “Infectious disease” was available for 1.8% of participants and “Metabolics” was available for 3.5% of participants.) |
| [Urine Assays](https://biobank.ndph.ox.ac.uk/showcase/label.cgi?id=100083) | -- | Variables within whole category | We used raw UKB variables. |
| [Physical Measures](https://biobank.ndph.ox.ac.uk/showcase/label.cgi?id=100006) |  | Variables within [Blood pressure](https://biobank.ndph.ox.ac.uk/showcase/label.cgi?id=100011), [Hearing test](https://biobank.ndph.ox.ac.uk/showcase/label.cgi?id=100049), [Arterial stiffness](https://biobank.ndph.ox.ac.uk/showcase/label.cgi?id=100007), [Hand grip strength](https://biobank.ndph.ox.ac.uk/showcase/label.cgi?id=100019), [Anthropometry](https://biobank.ndph.ox.ac.uk/showcase/label.cgi?id=100008), [Eye measures](https://biobank.ndph.ox.ac.uk/showcase/label.cgi?id=100013), [Bone-densitometry of heel](https://biobank.ndph.ox.ac.uk/showcase/label.cgi?id=100018), [Spirometry](https://biobank.ndph.ox.ac.uk/showcase/label.cgi?id=100020) | Admin fields (e.g. Carotid ultrasound measurement completed) and bulk data fields (e.g. [ECG datasets](https://biobank.ndph.ox.ac.uk/showcase/field.cgi?id=20205)) were excluded from input features.  Two ECG categories were excluded from input features: (i) “[ECG at rest, 12-lead](https://biobank.ndph.ox.ac.uk/showcase/label.cgi?id=104)”, due to unavailability at baseline, and (ii) “[ECG during exercise](https://biobank.ndph.ox.ac.uk/showcase/label.cgi?id=100012)”, because its analysis requires expert knowledge.  For sub-categories of eye measures, only 2 fields within “[Visual acuity](https://biobank.ndph.ox.ac.uk/showcase/label.cgi?id=100017)” were considered because UKB website indicates that “*For non-specialists, the primary items of interest are* [*Field 5201*](https://biobank.ndph.ox.ac.uk/showcase/field.cgi?id=5201) *and* [*Field 5208*](https://biobank.ndph.ox.ac.uk/showcase/field.cgi?id=5208)*.*” “[Autorefraction](https://biobank.ndph.ox.ac.uk/showcase/label.cgi?id=100014)” was not considered because it requires expert knowledge to analyse. “[Retinal optical coherence tomography](https://biobank.ndph.ox.ac.uk/showcase/label.cgi?id=100016)” and “[Intraocular pressure](https://biobank.ndph.ox.ac.uk/showcase/label.cgi?id=100015)” only contain data for 100k participants, hence were not considered.  Within the “[Spirometry](https://biobank.ndph.ox.ac.uk/showcase/label.cgi?id=100020)” category, we only included FVC, FEV1 and PEF measures, by taking the average value of multiple measurements at baseline. |
| [Imaging](https://biobank.ndph.ox.ac.uk/showcase/label.cgi?id=100003) | -- | -- | Not available at baseline. |
| [Cognitive function](https://biobank.ndph.ox.ac.uk/showcase/label.cgi?id=100026) | [Reaction time](https://biobank.ndph.ox.ac.uk/showcase/label.cgi?id=100032) | [Mean time to correctly identify matches](https://biobank.ndph.ox.ac.uk/showcase/field.cgi?id=20023) | We used the raw UKB variable. |
|  | [Numeric memory](https://biobank.ndph.ox.ac.uk/showcase/label.cgi?id=100029) | [Maximum digits remembered correctly](https://biobank.ndph.ox.ac.uk/showcase/field.cgi?id=4282) | Only this field was considered because UKB website indicates that “*For non-specialists, the primary item of interest is* [*Field 4282*](https://biobank.ndph.ox.ac.uk/showcase/field.cgi?id=4282).” |
|  | [Fluid intelligence](https://biobank.ndph.ox.ac.uk/showcase/label.cgi?id=100027) | [Fluid intelligence score](https://biobank.ndph.ox.ac.uk/showcase/field.cgi?id=20016) | Only this field was considered because UKB website states “For non-specialists, the primary item of interest is [Field 20016](https://biobank.ndph.ox.ac.uk/showcase/field.cgi?id=4282).” |
|  | [Trial making](https://biobank.ndph.ox.ac.uk/showcase/label.cgi?id=505),  [Matrix pattern completion](https://biobank.ndph.ox.ac.uk/showcase/label.cgi?id=501),  [Tower rearranging](https://biobank.ndph.ox.ac.uk/showcase/label.cgi?id=503),  [Picture vocabulary](https://biobank.ndph.ox.ac.uk/showcase/label.cgi?id=504),  [Symbol digit substitution](https://biobank.ndph.ox.ac.uk/showcase/label.cgi?id=502),  [Paired associate learning](https://biobank.ndph.ox.ac.uk/showcase/label.cgi?id=506) | -- | Only available from imaging visit. |
|  | [Prospective memory](https://biobank.ndph.ox.ac.uk/showcase/label.cgi?id=100031) | [Prospective memory result](https://biobank.ndph.ox.ac.uk/showcase/field.cgi?id=20018) | We used the raw UKB variable. |
|  | [Pairs matching](https://biobank.ndph.ox.ac.uk/showcase/label.cgi?id=100030) | -- | UKB website states “*For non-specialists, the primary item of interest is* [*Field 399*](https://biobank.ndph.ox.ac.uk/showcase/field.cgi?id=4282) *and (for the pilot) FID 10137*.” However this primary field was unavailable. |
|  | [Lights pattern memory](https://biobank.ndph.ox.ac.uk/showcase/label.cgi?id=100028),  [Word production](https://biobank.ndph.ox.ac.uk/showcase/label.cgi?id=100077) | -- | Both categories contain only pilot fields and hence were not used. |

### 3. Machine Learning (ML) Methods

#### 3.1 Hyper-parameter tuning of XGBoost

Parameters of the eXtreme Gradient Boosting (XGBoost) algorithm fall into two main categories: (i) tree-specific parameters that guide the structuring of individual trees at each step, and (ii) regularisation parameters that reduce the chance of over-fitting and enhance generalisation (full details in Supplementary Table 3).

The large set of tree-specific and regularisation parameters enables the construction of sophisticated models for better prediction, but adds difficulty in searching for the optimal parameters. Unlike classical statistical models whose parameters are directly estimated from the data, XGBoost machines usually require manual specification of parameters due to their complexity. Such parameters are often referred to as hyper-parameters, and the choice of their values has a direct impact on the model performance. Therefore, the process of searching for parameters that yield the optimal model (i.e. hyper-parameter tuning) is crucial for training an XGBoost machine.

Common strategies for hyper-parameter tuning include grid search ^23^, random search ^4^ , and Bayesian optimization ^5^. In this study, we applied grid search with 5-fold cross-validation (CV) on training data using Area Under the Receiver operating characteristic Curve (AUC) as the evaluation metric.

During the grid search, each combination of parameters specified in a grid was fitted to the model. For example, if one specifies a grid with depth of tree set to be {2, 3, 4} and number of trees to be {10, 20}, the grid search will fit the model with each of 6 possible combinations. The optimal combination was obtained by computing the average AUC obtained from the five validation sets and selecting the set with the highest value.

The drawback of this approach is its high computational cost for high-dimensional search spaces, which XGBoost is subject to due to its large set of parameters. Therefore, we further applied the following tuning strategy to improve the efficiency of the grid search.

We started with a relatively high learning rate of 0.1 to determine the corresponding approximate number of trees. We then tuned the tree-specific parameters followed by the tuning of regularisation parameters. Finally, we lowered the learning rate and increased the number of trees accordingly to obtain a more robust model (Supplementary Table 3).

Supplementary Table 3. Hyper-parameter tuning strategy of our XGBoost machine via grid search with 5-fold CV. We followed the naming convention of parameters by [Scikit-Learn API](https://xgboost.readthedocs.io/en/stable/python/python_api.html#module-xgboost.sklearn). max(AUC_CV_): Highest AUC obtained from 5-fold CV on training data.

| Steps | Search Ranges of parameters | Explanation |
| --- | --- | --- |
| Step 1: Build a baseline model | learning_rate$=$0.1 early_stopping_rounds$=$50 n_estimators$\in$ [1..500] | Learning rate (learning_rate) is inversely correlated with the number of trees (n_estimators). Lower learning rates typically yield better model performance, given a sufficient number of trees. However, computation becomes expensive with decreased learning rate and increased number of trees. Constructing a baseline model with a relatively high learning rate produces an approximate estimate of the number of trees required for a specific learning rate without sacrificing computational power.  Early stopping works as follows: for a pre-specified early stopping rounds (early_stopping_rounds) (e.g. 10), the model will train until there is no improvement in validation score in the next 10 rounds. This enables users to grow a complex model, minimise the chance of over-fitting, save computation time, and obtain a more accurate number of trees. |
| Step 2: Tune max_depth and min_child_weight | max_depth $\in$ [1..8]  min_child_weight$\in$ {1, 3, 5}  max(AUC_CV_)$=$0.662 | Parameter max_depth specifies the maximum depth of tree. The higher the value, the more complex the model. Parameter min_child_weight defines the minimum sum of weights of all observations required in a child. The higher the value, the more conservative the model. |
| Step 3: Tune gamma | gamma $\in$ {0, 0.2, 0.4, 0.6, 0.8, 1, 1.4, 1.6}  max(AUC_CV_)$=$0.662 | Parameter gamma is the minimum loss reduction required to make a split. The higher the value, the more conservative the model. |
| Step 4: Tune subsample and colsample_bytree | subsample$\in$ {0.6, 0.7, 0.8, 0.9}  colsample_bytree$\in${0.6,0.7,0.8,0.9}  max(AUC_CV_)$=$0.664 | Parameter subsample is the fraction of observations to be randomly sampled for each tree. Parameter colsample_bytree is the fraction of features to be randomly sampled for each tree. |
| Step 5: Tune regularisation parameters | lambda $\in$ {1e-5, 0.01, 0.1, 0.5, 1, 5, 8, 10, 12, 14, 16, 18, 20, 22, 100}  max(AUC_CV_)$=$0.665 | For regularisation parameters lambda and alpha, one has the option to choose which one to use for penalising weights of leaves: lambda corresponds to L2 regularisation term that encourages weights to be small, whereas alpha corresponds to L1 regularization that encourages weights to be 0. Here we took a more conservative approach for tuning lambda instead of alpha. |
| Step 6: Tune scale_pos_weight | scale_pos_weight $\in${1, 25, 100}  max(AUC_CV_)$=$0.665 | Parameter scale_pos_weight controls the balance of positive and negative weights, which can be useful for unbalanced classification (e.g. binary outcome that has substantially fewer cases than controls). |
| Step 7: Lower **learning_rate** and increase **n_estimators** | learning_rate $\in$ {0.1, 0.01, 0.001}  n_estimators$\in$[1..50000]  early_stopping_rounds=200  max(AUC_CV_)$=$0.668 | Here we lowered the learning rate (learning_rate) and increased the number of trees (n_estimators) for a more robust model. |

#### 3.2 SHAP values

SHapley Additive exPlanation (SHAP) values originated from game theory ^6^ as a metric to compute fair contribution among players. It has been recently incorporated into machine learning models where it is being developed to aid model interpretation. For the purpose of feature selection, SHAP values compute the marginal contribution of each feature to the model prediction among all possible coalitions, which takes account of interactions among features.

As a member of the additive feature attribution methods, it uses a simple explanation model to approximate the prediction from a complex machine learning model ^7^:

$$f\left( \boldsymbol{x} \right)=g\left( \boldsymbol{x}^{'} \right)= \emptyset_{0}+ \sum_{i=1}^{M} \emptyset_{i}\boldsymbol{x}_{i}^{'}$$

(1)

Where $\boldsymbol{x}$ is the original input feature, $\boldsymbol{x}^{\boldsymbol{'}}$ is the simplified input, mapped through a function where $\boldsymbol{x}=h_{x}(\boldsymbol{x}^{\boldsymbol{'}})$, $f\left( \boldsymbol{x} \right)$ is the model output that is computed by default in log odds scale for binary classifications, $\emptyset_{0}$ is the output when no input is present, *M* is the total number of features, and $\emptyset_{i}\mathbb{\in R}$ is the attribution value (i.e. SHAP value) of feature $i$ ^7^.

The SHAP value of feature $i$ is computed as follows: for each possible subset of features $S\subseteq F\backslash\{i\}$ (where $F$ represents the set of all features), two models are trained incorporating the potential dependency between features: one including feature $i$ and one without it. The difference between the outputs of the two models is then computed as $f_{S\cup\{i\}}\left( \boldsymbol{x}_{S\cup\{i\}} \right)- f_{S}\left( \boldsymbol{x}_{S} \right)$, where $\boldsymbol{x}_{S}$ indicates the values of input features in the subset $S$. $\emptyset_{i}$ is computed as the weighted average of all possible differences, where the weight captures the feature space of each subset, shown in Equation (2),

$$\emptyset_{i}= \sum_{S\subseteq F\backslash\{i\}} \frac{\left| S \right|!\left( M-\left| S \right|-1 \right)!}{M!} \left[ f_{S\cup\{i\}}\left( \boldsymbol{x}_{S\cup\{i\}} \right)- f_{S}\left( \boldsymbol{x}_{S} \right) \right]$$

(2)

where $F$ is the set of all features. $\left| S \right|!$ is the number of features in $S$ where $S\subseteq F\backslash\{i\}$ represents all the possible subsets without feature $i$.

A major challenge for computing SHAP values is that the computation cost grows exponentially as the number of features increases. This is resolved by computing SHAP values locally (i.e. each feature attribution is computed using one sample) ^8^. For tree-based ML models, the internal tree structure is utilised for faster computation.

More specifically, for a dataset with $N$ samples and $M$ features (i.e. $N\times M$), SHAP values are generated in the same dimension where the attribution of feature $i$, $\emptyset_{ij}$, is computed locally using each sample for individual $j$. In the summary bar plot (e.g. Supplementary Figure 1), the mean absolute SHAP (SHAP_ma_) value of each feature is aggregated by taking the mean over all samples:

$${\mathrm{SHAP}\mathrm{ma}}_{i}= \frac{1}{N} \sum_{j=1}^{N} |\emptyset_{ij}|$$

In addition to SHAP values, we also implemented two different feature importance methods: XGBoost default feature importance (“weight”) and permutation based feature importance. We observed inconsistency between the XGBoost default feature importance (“weight”) ranking and the SHAP values ranking. The permutation based method required high computation cost compared with the others, and due to high collinearity between features it did not produce feasible results for further inspection.

#### 3.3 HistGBM

As a sensitivity analysis, we further investigated the robustness of feature ranking by exploring another ML method, the Histogram-based Gradient Boost Machines (HistGBM), inspired by LightGBM ^9^. Traditional GBM and HistGBM select the split points differently during the construction of trees. Traditional GBM sorts values of each continuous feature, and considers the average of each pair of adjacent values as splitting points when building trees, which can be computationally expensive for high-dimensional data. HistGBM improves the computational efficiency by discretising (i.e. binning) the continuous features to construct feature histograms (255 bins by default) during training, which significantly decreases the number of splitting points. This approximation often has little impact on model performance but dramatically reduces the memory consumption and accelerates the training speed. Besides faster implementation for large datasets, HistGBM also handles missing data by default (in the same way as XGBoost) and utilises L2 regularization to reduce over-fitting.

We adapted the same hyper-parameter tuning process (grid search with 5-fold CV) as in our XGBoost machine, except that HistGBM has fewer parameters to tune (Supplementary Table 4).

Supplementary Table 4. Hyper-parameter tuning strategy of HistGBM via grid search with CV. We followed the naming convention of parameters by [Scikit-Learn API](https://scikit-learn.org/stable/modules/generated/sklearn.ensemble.HistGradientBoostingClassifier.html). max(AUC_CV_): Highest AUC obtained from 5-fold CV on training data.

| Steps | Search Ranges of parameters |
| --- | --- |
| Step 1: Build a baseline model | learning_rate$=$0.1  n_iter_no_change*$=$50  max_iter* $\in$[1..500] |
| Step 2: Tune tree-specific parameters | max_dpeth $\in$ [1..8]  min_samples_leaf* $\in${20, 30,40,50,60,70}  max(AUC_CV_)$=$ 0.663 |
| Step 3: Tune L2 regularisation parameter | lambda $\in${1e-6, 1e-5, 5e-5, 0, 0.01, 0, 0.1, 1, 5, 10, 20, ,25, 30, 35, 40, 45, 50, 100}  max(AUC_CV_)$=$0.664 |
| Step 4: Lower **learning_rate** and increase **max_iter** | learning_rate $\in$ {0.1, 0.01, 0.001}  n_estimators$\in$[1..50000]  early_stopping_rounds$=$200  max(AUC_CV_)$=$0.664 |

*Note*:* n_iter_no_change *controls when to early stop (analogous to* early_stopping_rounds *in XGBoost);* max_iter *represents the number of trees;* min_samples_leaf *defines minimum samples per leaf.*

During Step 4 of hyper-parameter tuning, we observed the model with learning_rate$=$0.001 had the highest AUC; however, the improvement compared to model with learning_rate$=$0.01 was modest. After taking computation time into account, the optimal set of hyper-parameters for HistGBM were found to be: learning_rate $=$ 0.01, max_iter $=$ 1,366, max_depth $=$ 2, min_samples_leaf $=$ 30, lambda $=$ 35. The AUC obtained from the 5-fold CV on training data was 0.664 and the AUC on test data was 0.663.

The top 20 features with SHAP values from HistGBM are shown in Supplementary Figure 1. We observed a high level of overlap with those from XGBoost (Figure 4 in manuscript), which confirms the robustness of our risk factor discovery.

The six new features not selected by XGBoost are:

- Physical activity features (Duration walking for pleasure, Summed MET minutes per week for all activity)
- Hormone (SHBG)
- Anthropometry (Trunk fat-free mass)
- Others (Country of birth (UK/elsewhere), UK country of residence).

Three of these six were on the borderline of being included in the top 20 by XGBoost and SHAP values; they are: trunk fat-free mass (ranked 21), summed MET minutes per week for all activity (ranked 23), and SHBG (ranked 29).

Although duration of walking for pleasure was ranked 10 by HistGBM (but only 135 by XGBoost), it may have relatively little impact since the more detailed physical activity feature, summed MET minutes per week for all activity, was already ranked among the top 20 features by HistGBM. Variables in the "Others" category could be due to the artefacts of censoring, e.g. the short follow-up time in Wales may have contributed to fewer breast cancer cases there.

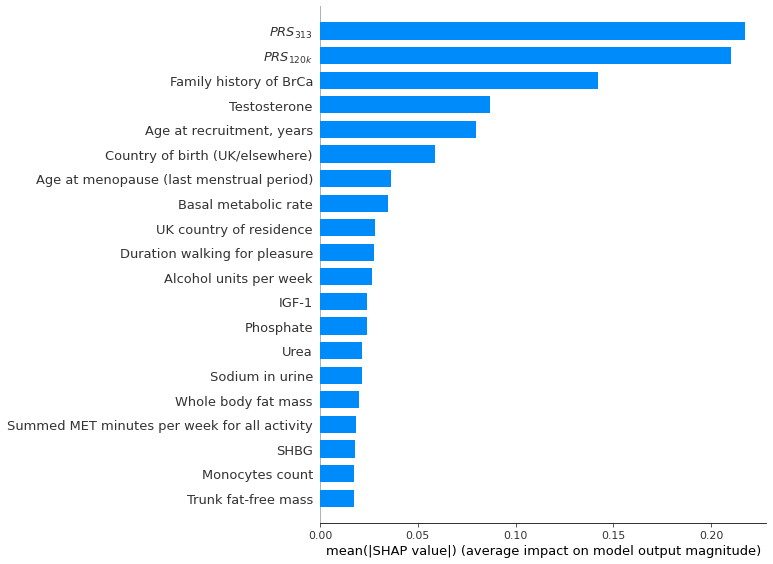

Supplementary Figure 1. The top 20 most important features for the risk of breast cancer (BrCa), according to the HistGBM and SHAP values. Noticeably, both BrCa PRS are deemed of much higher importance than the remaining phenotypic features. BrCa: Breast Cancer. SHAP: SHapley Additive explanation. mean(|SHAP value|): Mean absolute SHAP value.

### 4. Tables

#### 4.1 Full Cox model output

Supplementary Table 5. Cox regression of time until breast cancer in post-menopausal women (3,252 events). Baseline model represents multivariable Cox model containing two PRS and established risk factors. New model represents multivariable Cox model containing all covariates in baseline model and novel features discovered by XGBoost.

|  | Baseline model | | | New model | | |
| --- | --- | --- | --- | --- | --- | --- |
| Coefficient | HR | 95% CI | p | HR | 95% CI | p |
| $PRS_{120k}$ **quintiles** |  |  |  |  |  |  |
| Q1: Lowest score | 1 |  |  | 1 |  |  |
| Q2 | 1.36 | (1.18, 1.58) | <0.01 | 1.36 | (1.17, 1.58) | <0.01 |
| Q3 | 1.53 | (1.32, 1.77) | <0.01 | 1.53 | (1.32, 1.77) | <0.01 |
| Q4 | 1.72 | (1.49, 2.00) | <0.01 | 1.72 | (1.48, 1.99) | <0.01 |
| Q5: Highest score | 2.34 | (2.02, 2.72) | <0.01 | 2.33 | (2.01, 2.70) | <0.01 |
| $PRS_{313}$ **quintiles** |  |  |  |  |  |  |
| Q1: Lowest score | 1 |  |  | 1 |  |  |
| Q2 | 1.17 | (1.01, 1.35) | 0.04 | 1.17 | (1.01, 1.36) | 0.04 |
| Q3 | 1.40 | (1.21, 1.61) | <0.01 | 1.40 | (1.21, 1.62) | <0.01 |
| Q4 | 1.67 | (1.45, 1.93) | <0.01 | 1.68 | (1.45, 1.94) | <0.01 |
| Q5: Highest score | 2.20 | (1.90, 2.54) | <0.01 | 2.21 | (1.91, 2.56) | <0.01 |
| **Age at recruitment, years** | 1.03 | (1.02, 1.04) | <0.01 | 1.03 | (1.03, 1.04) | <0.01 |
| **Whole body fat mass** | 1.01 | (1.01, 1.02) | <0.01 | 1.00 | (0.99, 1.01) | 0.92 |
| **Age at menopause (last menstrual period)** | 1.02 | (1.01, 1.02) | <0.01 | 1.02 | (1.01, 1.02) | <0.01 |
| **Daily alcohol intake** | 1.06 | (1.04, 1.09) | <0.01 | 1.07 | (1.04, 1.09) | <0.01 |
| **Age at first birth (Categorical)** |  |  |  |  |  |  |
| No Births | 1 |  |  | 1 |  |  |
| <20 | 1.05 | (0.86, 1.29) | 0.61 | 1.08 | (0.89, 1.33) | 0.44 |
| 20-30 | 1.07 | (0.93, 1.24) | 0.34 | 1.10 | (0.95, 1.27) | 0.21 |
| 30-40 | 1.24 | (1.07, 1.43) | <0.01 | 1.24 | (1.07, 1.44) | <0.01 |
| >=40 | 1.09 | (0.72, 1.67) | 0.67 | 1.09 | (0.72, 1.66) | 0.68 |
| **Family history of BrCa** |  |  |  |  |  |  |
| No family history of BrCa | 1 |  |  | 1 |  |  |
| Family history of BrCa | 1.34 | (1.22, 1.47) | <0.01 | 1.33 | (1.21, 1.46) | <0.01 |
| **Summed MET minutes per week for all activity** | 1.00 | (1.00, 1.00) | 0.07 | 1.00 | (1.00, 1.00) | 0.06 |
| **G03FAuser at baseline** | 1.90 | (1.58, 2.27) | <0.01 | 1.70 | (1.41, 2.04) | <0.01 |
| **Age when periods started (menarche)** | 0.98 | (0.96, 1.01) | 0.17 | 0.98 | (0.96, 1.01) | 0.17 |
| **IGF-1** | 1.01 | (1.00, 1.02) | <0.01 | 1.01 | (1.01, 1.02) | <0.01 |
| **Testosterone** | 1.12 | (1.08, 1.16) | <0.01 | 1.12 | (1.08, 1.16) | <0.01 |
| **Number of live births** | 0.94 | (0.90, 0.99) | 0.02 | 0.94 | (0.89, 0.98) | <0.01 |
| **Urea** |  |  |  | 0.95 | (0.92, 0.98) | <0.01 |
| **Basal metabolic rate** |  |  |  | 1.00 | (1.00, 1.00) | <0.01 |
| **Phosphate** |  |  |  | 0.67 | (0.52, 0.88) | <0.01 |
| **Sodium in urine** |  |  |  | 1.00 | (1.00, 1.00) | 0.11 |
| **Red Blood Cell Count** |  |  |  | 1.20 | (1.08, 1.34) | <0.01 |
| **Aspartate aminotransferase** |  |  |  | 1.00 | (0.99, 1.00) | 0.29 |
| **Creatinine (enzymatic) in urine** |  |  |  | 1.00 | (1.00, 1.00) | <0.01 |
| **Monocytes count** |  |  |  | 1.10 | (0.94, 1.28) | 0.22 |
| **Alkaline phosphatase** |  |  |  | 1.00 | (1.00, 1.00) | 0.12 |
| **C-reactive protein** |  |  |  | 1.00 | (0.99, 1.01) | 0.75 |

*BrCa, Breast Cancer; HR, hazard ratio (for continuous variables, HR represents each 1 standard deviation increase); CI, confidence interval; p, p-value. Note*: Baseline model contains all established BrCa risk factors whereas New model contains established BrCa risk factors and novel features from machine learning. Both PRS were categorised into quintiles; alcohol intake was scaled from weekly intake to daily intake for interpretation and direct comparison with literature; genetic array and first 10 PCs were adjusted in the model but omitted from the table. HR (95% CIs) of basal metabolic rate, sodium in urine, creatinine in urine, alkaline phosphatase and summed MET minutes per week for all activity from new model were presented as (1.00, 1.00) due to rounding.*

### 5. Plots

#### 5.1 Forest plot after excluding first 2 years follow-up

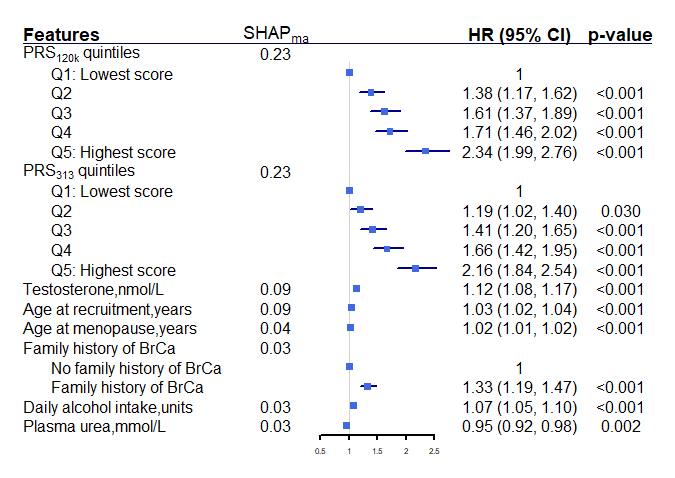

(a)

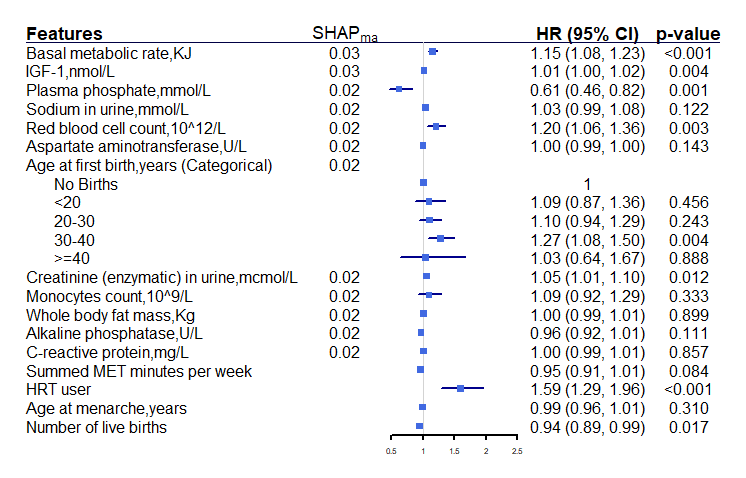

(b)

Supplementary Figure 2. Results obtained from our final multivariable Cox model after excluding first two years of follow-up, separated into two subplots for easy of reading: (a) top 8 features ranked by SHAP_ma_, (b) the remaining 16 features, whose bottom four features are established risk factors that are outside the top 20 features by SHAP_ma_. Both PRS were categorised into quintiles, alcohol intake was scaled from weekly intake to daily intake for easy interpretation and direct comparison with existing literature. Basal metabolic rate, sodium in urine, creatinine in urine, alkaline phosphatase, and summed MET minutes per week were standardised using the mean and standard deviation within each imputed dataset, hence the corresponding HR represents per 1 standard deviation increase. For other continuous variables, HR represents per 1 unit increase. Genetic array and first 10 PCs were adjusted in the model but omitted from the figure. SHAP: SHapley Additive explanation. SHAP_ma_: mean absolute SHAP value. BrCa: Breast Cancer. HR: hazard ratio. CI: confidence interval. HRT: hormone replacement therapy. MET: Metabolic Equivalent Task. U/L: units per litre.

#### 5.2 SHAP dependence plots

Our SHAP dependence plots not only revealed nonlinear relationships of phenotypic features with breast cancer, but also indicated potential interactions between PRS and a range of phenotypic features (e.g. age, family history, sodium in urine, and whole body fat mass).

| 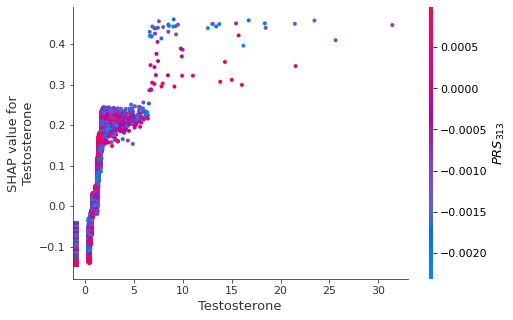  (a) | 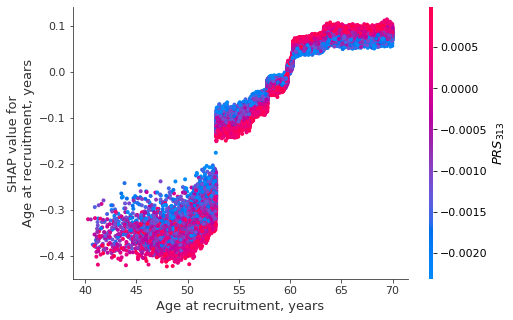  (b) |
| --- | --- |
| 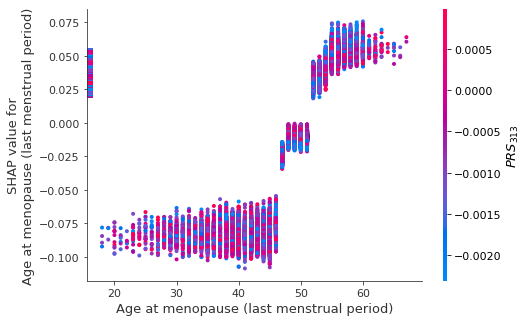  (c) | 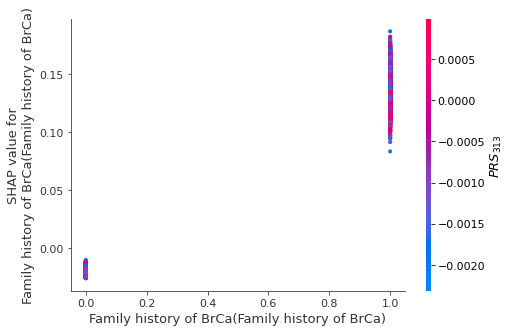  (d) |
| 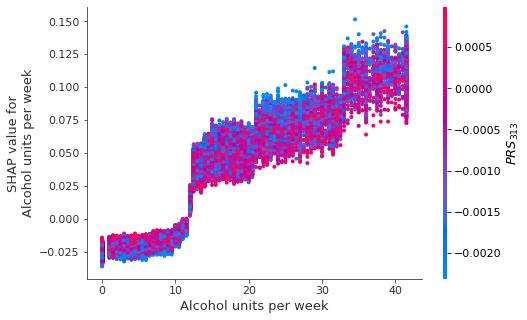  (e) | 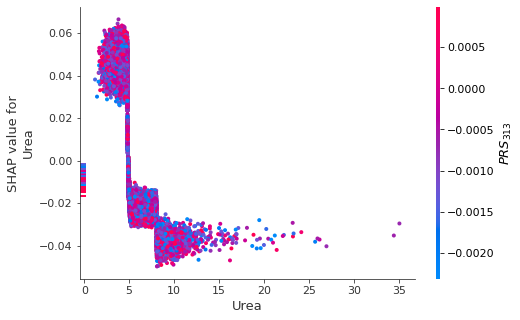  (f) |
| 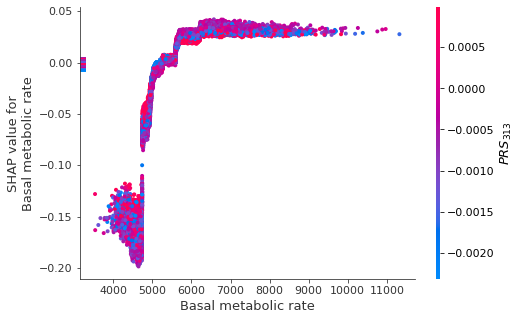  (g) | 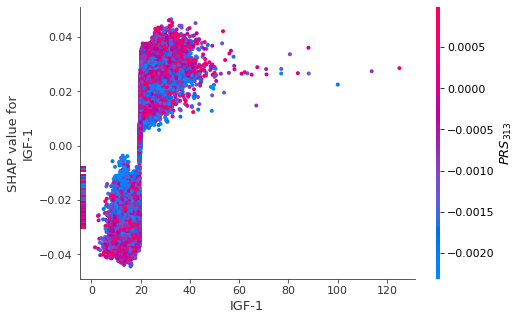  (h) |
| 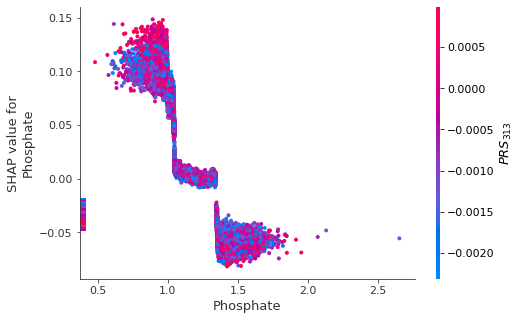  (i) | 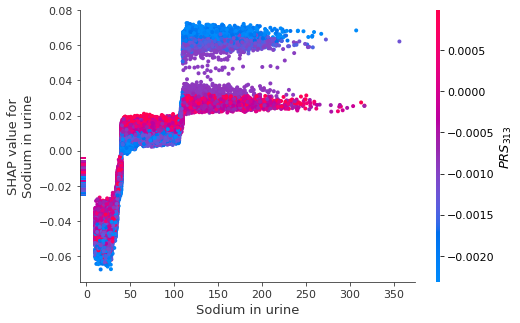  (j) |
| 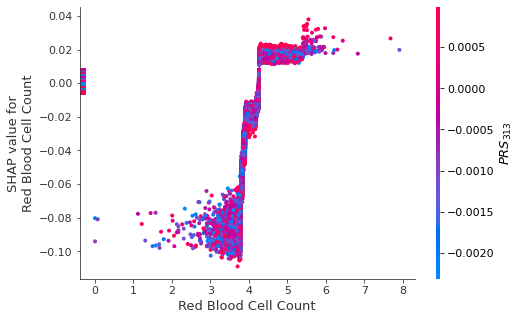  (k) | 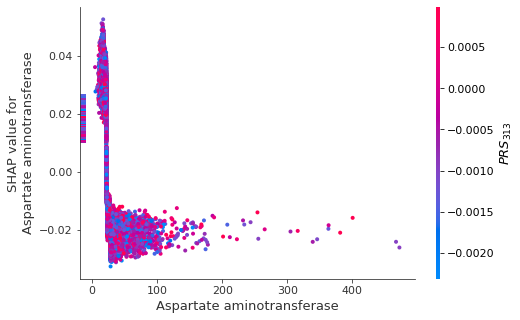  (l) |
| 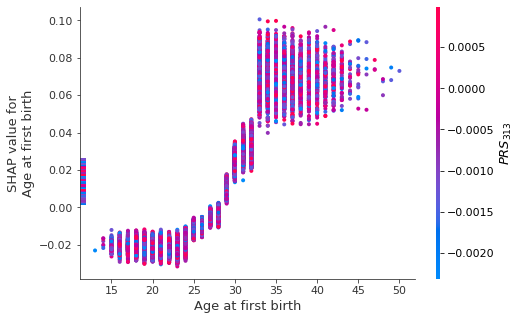  (m) | 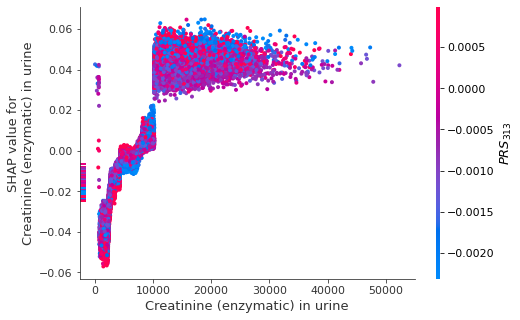  (n) |
| 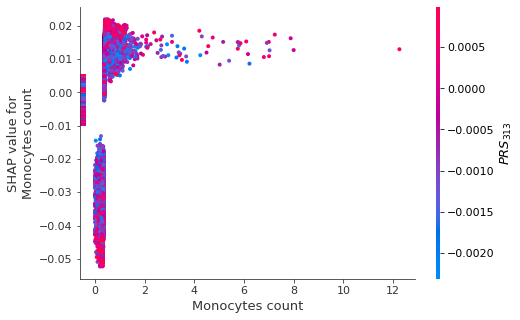 (o) | 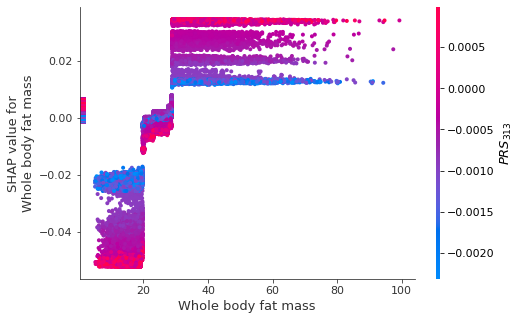  (p) |
| 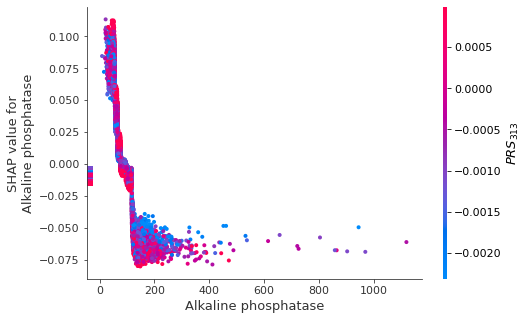  (q) | 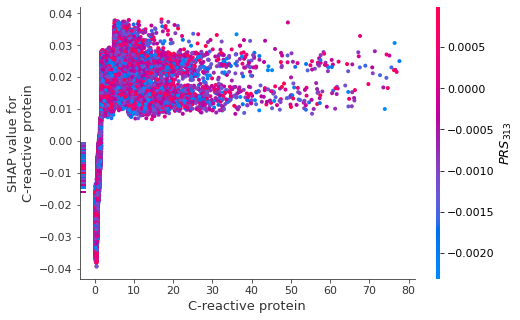  (r) |

*Supplementary Figure 3. SHAP dependence plots of the top 20 phenotypic features against PRS_313_ for breast cancer. Note: The vertical block of dots attached on the left y-axis represent individuals who had missing values of corresponding feature shown in x-axis. For example, in sub-figure (a), dots attached to the left y-axis with SHAP value for testosterone within [ -0.15, -0.05] are individuals with missing testosterone instead of having negative testosterone values.*

| 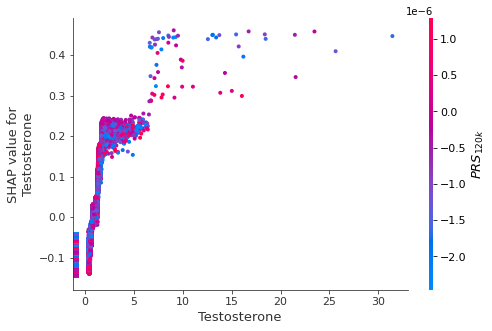  (a) | 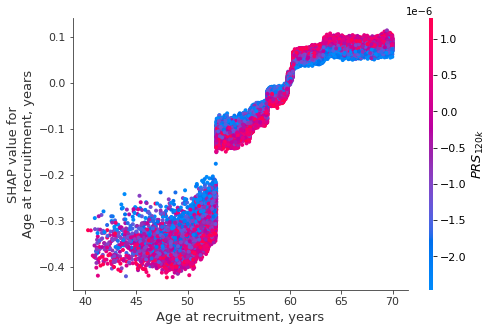  (b) |
| --- | --- |
| 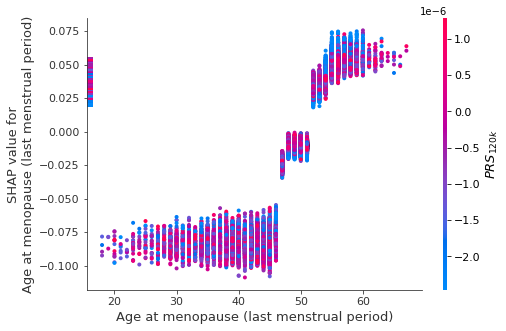 (c) | 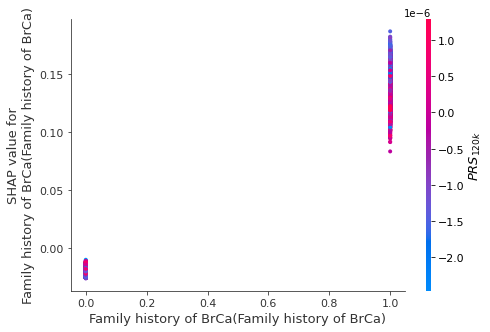  (d) |
| 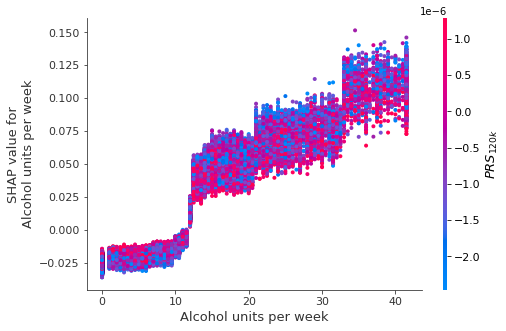 (e) | 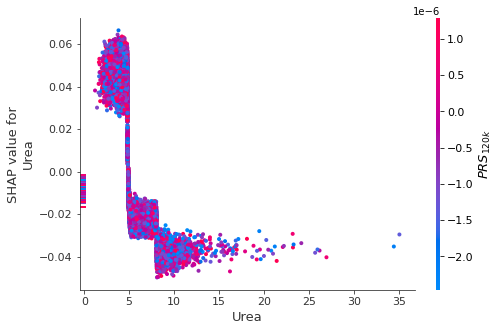  (f) |
| 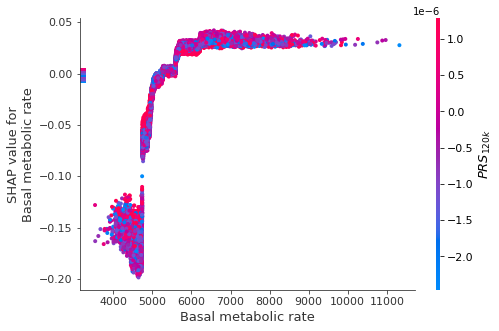  (g) | 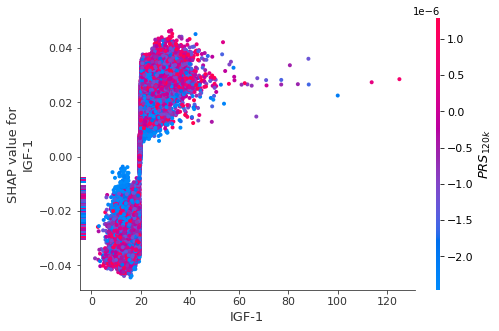  (h) |
| 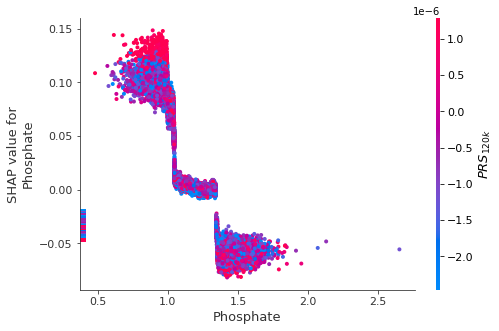 (i) | 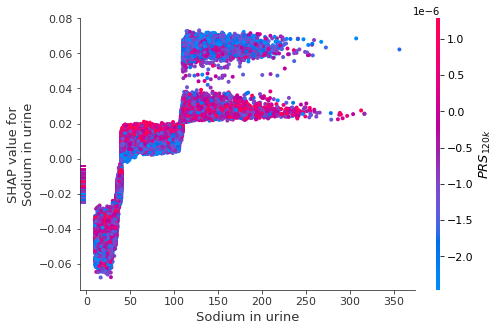  (j) |
| 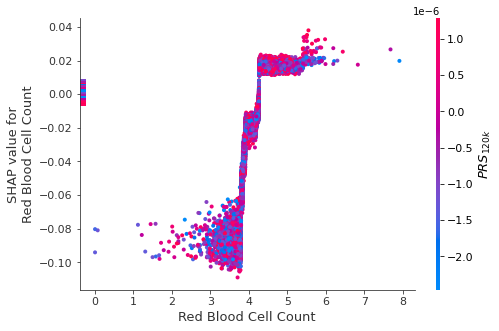 (k) | 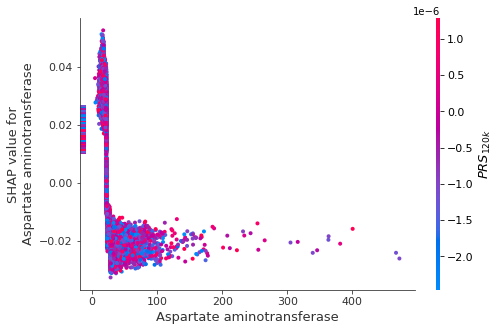  (l) |
| 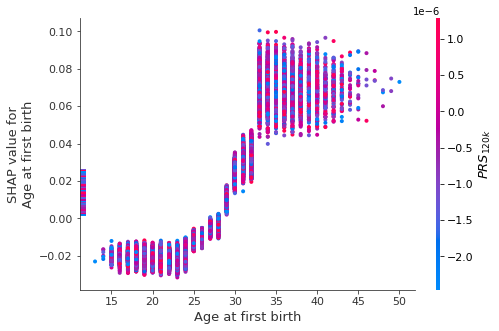  (m) | 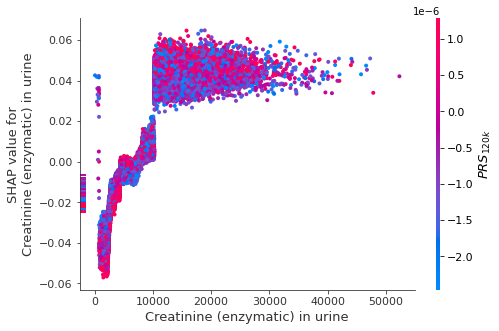  (n) |
| 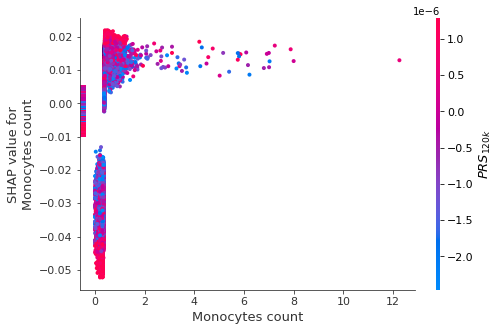 (o) | 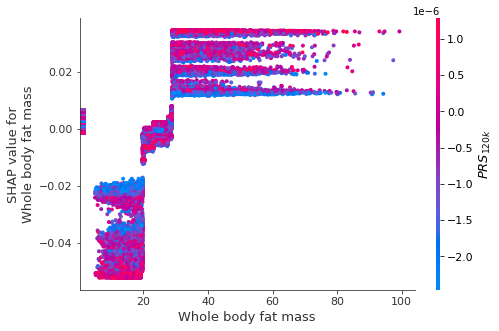 (p) |
| 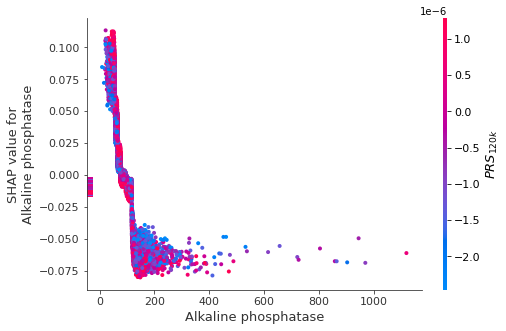  (q) | 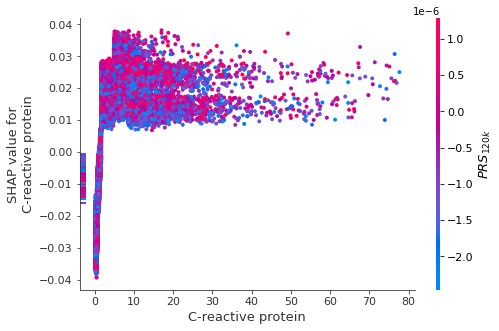  (r) |

*Supplementary Figure 4. SHAP dependence plots of the top 20 phenotypic features against PRS_120k_ for breast cancer. Note: The vertical block of dots attached on the left y-axis represent individuals who had missing values of corresponding feature shown in x-axis. For example, in sub-figure (a), dots attached to the left y-axis with SHAP value for testosterone within [ -0.15, -0.05] are individuals with missing testosterone instead of having negative testosterone values.*

### 6. SHAP dependence plot of age against PRS

We utilised SHAP dependence plots to identify potential interactions between PRS and potential risk factors, and subsequently constructed Cox models for in-depth investigation.

For both PRS_313_ and PRS_120k,_ SHAP dependence plots indicated potential effect modification of PRS with age, family history, sodium in urine and whole body fat mass (Supplementary Figure 3-4). We illustrate this in Supplementary Figure 5 which shows one of the most distinctive interactions, Age $\times$ PRS, where higher SHAP values indicate higher odds of developing breast cancer due to age. The PRS values are colour-coded from blue to red, representing PRS values from low to high.

| 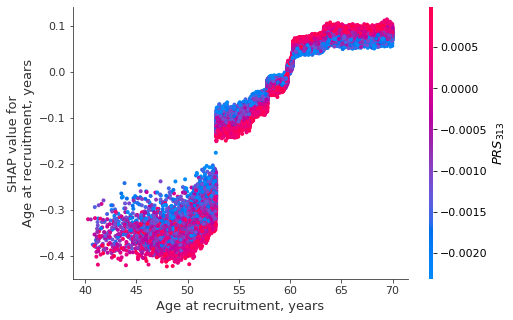(a) | (b)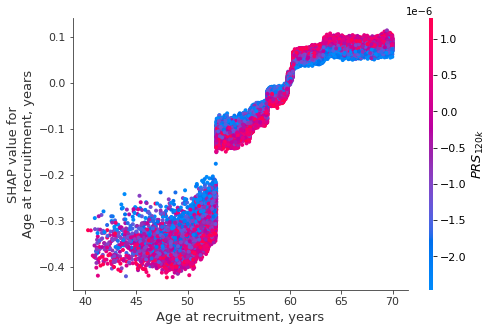 |
| --- | --- |

Supplementary Figure 5. SHAP dependence plot showing age against PRS, where each dot represents each participant in the study population; (a) PRS_313_ and (b) PRS_120k_. The y-axis shows the SHAP values attributed to age at recruitment (x-axis). Higher SHAP values indicate higher log(odds of developing breast cancer). The corresponding PRS value of each individual is represented by colour, with blue indicating low and red indicating high PRS values.

Both subplots in Supplementary Figure 5 show a clear non-linear main effect of age on developing breast cancer where the odds of developing breast cancer increase with age and plateau at approximately 62 years old. Similar patterns have been previously observed and referred to as Clemmensen's hook ^10^. When taking PRS (represented by colour) into account, we observed a vertical dispersion of colours clustered at three age groups: 40-53, 53-62, and 62-70 years. Such dispersion of colours suggests that PRS has an impact on the importance of age (as quantified by SHAP values). For example, women with a low PRS (blue) typically had higher SHAP values for age than those with a high PRS (red) among younger age groups (40-53 and 53-62 years), indicating higher risk of breast cancer at lower PRS value (i.e. higher PRS dampens the effect of age on breast cancer). By contrast, the higher relative location of the red dots above the blue for age group 62-70 years indicates higher risk of breast cancer at higher PRS value (while the effect of age on breast cancer plateaus for this age group).

The youngest age group of 40-53 years corresponds to the sparsest dots in Supplementary Figure 5, because menopause is relatively rare among this youngest age group. The substantial increase in SHAP values above 53 years of age corresponds to the local pattern in our dataset where a subgroup of people had an increasing shift in PRS values at age 53.

The unexpected patterns arose from Supplementary Figure 5 include (i) both PRS appeared to be “protective” for younger women, and the opposite for older women; (ii) the sudden increase (i.e. “jump”) of the SHAP value for age (SHAP_age_) at around age of 53 years. In the next section, we provided our investigation and explanation on the unusual patterns.

#### 6.1 Investigation in Cox models

As an additional exploratory analysis to explore potential interactions indicated by the SHAP dependence plots, we added “Age $\times$ PRS” interaction terms in our final multivariable Cox model. No significant effect modification of age by PRS was found ($p=0.14$ or Age $\times$ PRS_313_, $p=0.91$ for Age $\times$ PRS_120k_), but a marginal effect plot of age by PRS (Supplementary Figure 6) suggests that the effect of older age is slightly stronger in the highest PRS quintile compared to the lowest quintile. To explore further, we constructed three separate Cox models stratifying by age group: 40-53, 53-62, and 62-70 years. Our results showed higher risk of breast cancer at higher PRS values in all age groups (Supplementary Figure 7).

| 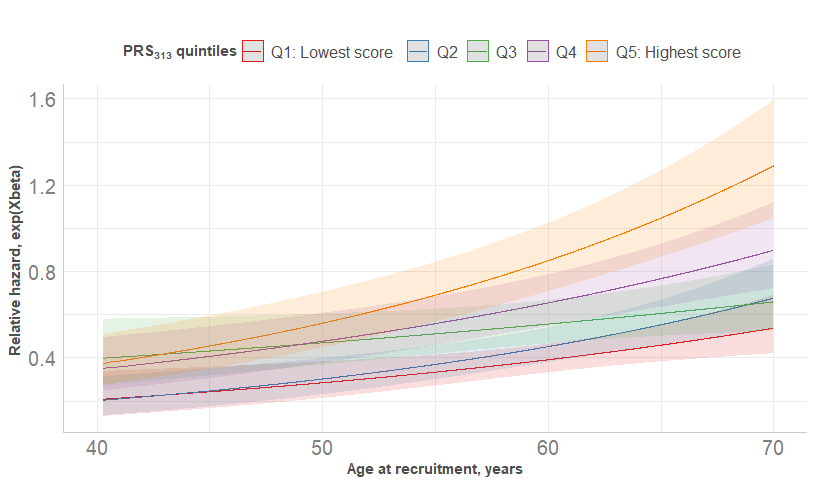  (a) | 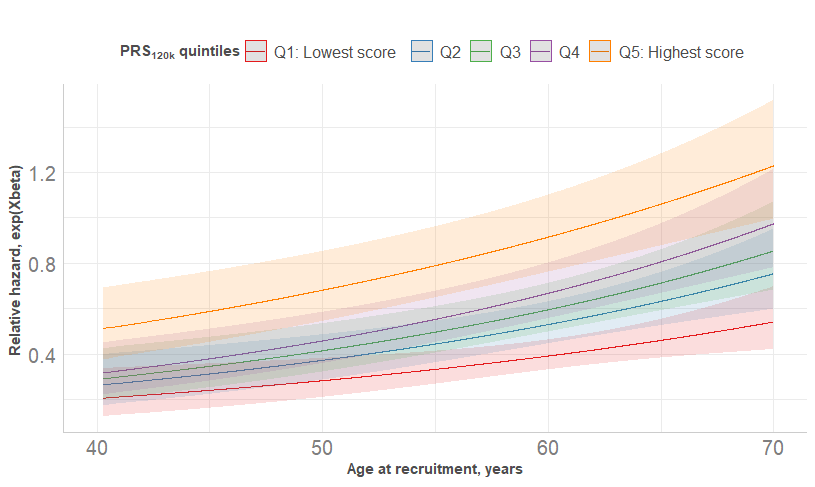  (b) |
| --- | --- |

Supplementary Figure 6. Marginal effect of age on relative hazard with pointwise 95% CI obtained from multivariable Cox model. Other continuous variables were kept as sample mean while categorical variables were kept at reference level.

| 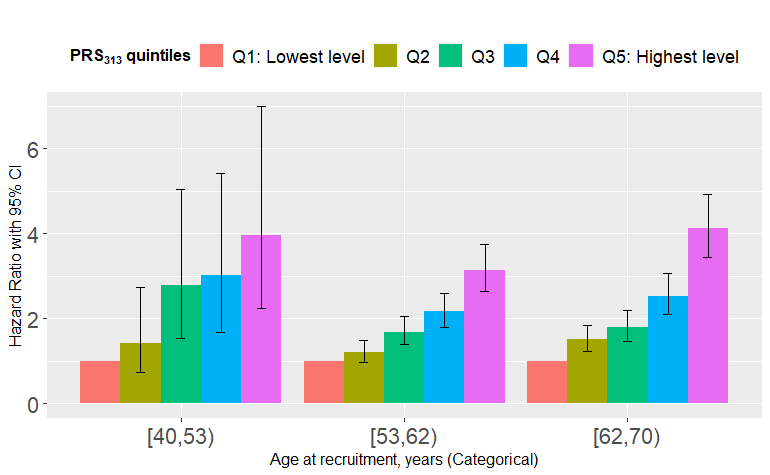  (a) | 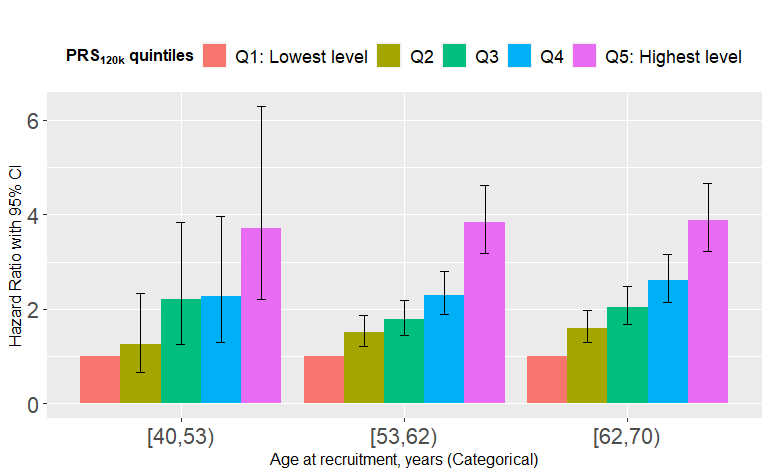  (b) |
| --- | --- |

Supplementary Figure 7. Hazard ratio with 95% confidence interval of PRS obtained from three separate Cox models stratified by age groups: [40,53), [53,62), [62,70) years, with adjustment of genetic array and first 10 PCs; (a) PRS_313_ and (b) PRS_120k_. First quintile of PRS, “Q1: Lowest level” was regarded as the reference level for comparison across other quintiles within each age group.

In summary, the interaction of Age $\times$ PRS was observed in the SHAP dependence plots, but it was not statistically significant in the subsequent Cox models.

#### 6.2 Investigation of the “jump” of SHAP values at “Age” of 53 years

The SHAP dependence plot (Supplementary Figure 5) of age against PRS shows a sudden increase (i.e. “jump”) of the SHAP value for age (${SHAP}_{age}$) at around age of 53 years, corresponding to a “jump” in the odds of developing breast cancer at age of 53 years. In this section we explore the potential reasons for such an unusual pattern.

##### 6.2.1 Step 1: The tree structure

We first checked our tree structures because tree-based models (e.g. XGBoost) are characteristically discontinuous. More specifically, we suspected some trees in our XGBoost model used condition “Age at recruitment $>$ 53 years” as a split point; that is, if participant had age$>$53, they would be assigned to the left child node, otherwise to the right child node. This will directly impact the SHAP values whose computation utilises information from the tree structure such as the number of samples under each leaf node.

Among the 1,571 trees in our XGBoost machine, 214 trees used “Age at recruitment” to split nodes with varying splitting points (median=57.8, IQR= 52.8-60.1 years) (Supplementary Figure 8).

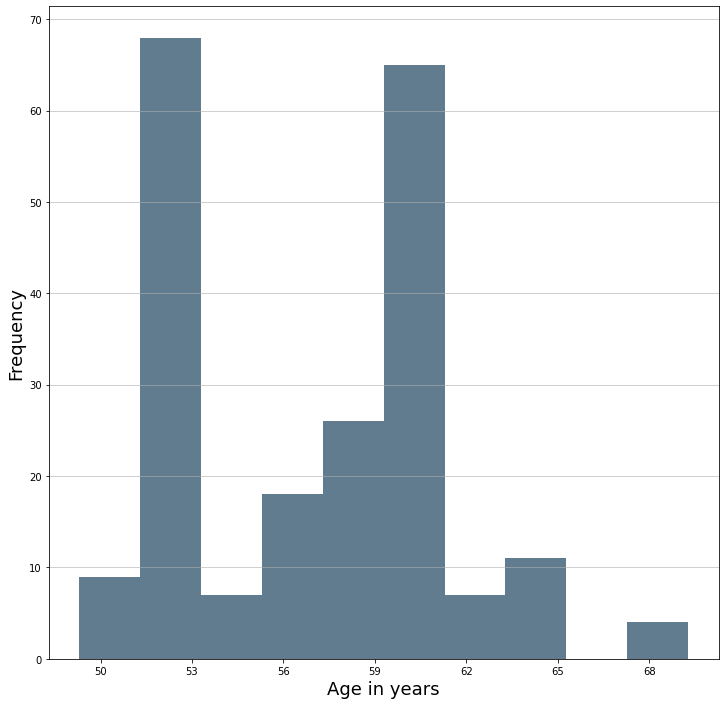

Supplementary Figure 8. Histogram of “Age at recruitment” used for splitting in the internal nodes of our XGBoost model.

Although “Age at recruitment” around 53 years old was frequently used as a splitting point, the obvious variation in the histogram of the splitting points indicates that the tree structure does not constitute enough evidence to explain the “jump”.

##### 6.2.2 Step 2: Descriptive statistics

Next, we compared baseline characteristics of training data between individuals of Age $<$53 years and those of Age $\geq$ 53 years (Supplementary Table 6). The aim is to detect whether there is any substantial difference across age of 53 years in these characteristics that might contribute to the jump. The baseline characteristics appear balanced except that the “Age $<$ 53 years” group had higher creatinine in urine and more HRT users.

Supplementary Table 6. Baseline characteristics of the training data by whether age below 53 years old. Median (interquartile ranges, IQR) are presented for continuous variables, while frequency (percentage) are reported for categorical variables. Note*: Both PRS were multiplied by their number of alleles for easy comparison. BrCa: Breast Cancer. HRT: hormone replacement therapy.

|  | Age $<$53 (N=6603) | Age $\geq$ 53 (N=76847) |
| --- | --- | --- |
| $PRS_{120k}$* | -0.14 (-0.31, 0.03) | -0.14 (-0.31, 0.04) |
| $PRS_{313}$* | -0.41 (-0.82, 0.01) | -0.41 (-0.82, 0.00) |
| **Testosterone, nmol/L** | 1.04 (0.74, 1.41) | 0.97 (0.69, 1.32) |
| **Age at recruitment, years** | 50.97 (49.28, 51.94) | 61.82 (58.28, 65.25) |
| **Age at menopause (last menstrual period), years** | 48.00 (44.00, 50.00) | 51.00 (48.00, 53.00) |
| **Alcohol units per week** | 6.00 (0.00, 13.50) | 6.00 (0.00, 12.00) |
| **Plasma urea, mmol/L** | 4.94 (4.25, 5.74) | 5.38 (4.64, 6.22) |
| **Basal metabolic rate, KJ** | 5594.00 (5230.00, 6025.00) | 5485.00 (5138.00, 5908.00) |
| **IGF-1, nmol/L** | 21.66 (17.90, 25.21) | 20.05 (16.47, 23.60) |
| **Plasma phosphate, mmol/L** | 1.22 (1.12, 1.32) | 1.21 (1.12, 1.30) |
| **Sodium in urine, millimole/L** | 59.70 (36.80, 91.60) | 55.10 (35.50, 83.20) |
| **Red Blood Cell Count, 10^12 cells/Litre** | 4.30 (4.09, 4.51) | 4.34 (4.13, 4.56) |
| **Aspartate aminotransferase, U/L** | 22.90 (20.00, 26.70) | 23.90 (20.90, 27.60) |
| **Age at first birth, years** | 26.00 (23.00, 30.00) | 25.00 (22.00, 28.00) |
| **Creatinine (enzymatic) in urine, micromole/L** | 5746.50 (3424.75, 9676.25) | 5523.00 (3418.00, 8993.00) |
| **Monocytes count, 10^9 cells/Litre** | 0.40 (0.30, 0.50) | 0.41 (0.33, 0.51) |
| **Whole body fat mass, Kg** | 24.60 (19.10, 31.50) | 25.60 (20.40, 32.10) |
| **Alkaline phosphatase, U/L** | 83.80 (69.70, 100.20) | 86.80 (73.50, 102.50) |
| **C-reactive protein, mg/L** | 1.24 (0.59, 2.83) | 1.44 (0.71, 2.97) |
| **Family history of BrCa** | 650 (9.8%) | 8479 (11.0%) |
| **Summed MET minutes per week for all activity** | 1706.00 (788.00, 3439.50) | 1786.00 (834.00, 3546.00) |
| **HRT user** | 342 (5.2%) | 1600 (2.1%) |
| **Age when periods started (menarche)** | 13.00 (12.00, 14.00) | 13.00 (12.00, 14.00) |
| **Number of live births** | 2.00 (0.00, 2.00) | 2.00 (1.00, 2.00) |

##### 6.2.3 Step 3: Local explanations

Since baseline characteristics did not reveal any distinctive difference between Age $<$53 and Age $\geq$ 53 years, we zoomed into the local region of the jump where 35 individuals had ${SHAP}_{age} \in(-0.21,-0.14)$ to further investigate how ${SHAP}_{age}$ was computed for each of them. As mentioned in Section 3.2, SHAP values are computed using model output $f\left( \boldsymbol{x} \right)$ which can be visualised locally (i.e. how much features have influenced the model output for each sample) using force plots.

Hence the investigation was carried out by visualising and comparing the model predictions of these 35 individuals. For the purpose of demonstration, we showed the force plots of three representative samples with ${SHAP}_{age}=-0.21$ (i.e. lower bound: before the jump at age 53), ${SHAP}_{age}=-0.175$ (i.e. middle point), ${SHAP}_{age}=-0.14$ (i.e. upper bound: after the jump at age 53) (Supplementary Figure 9).

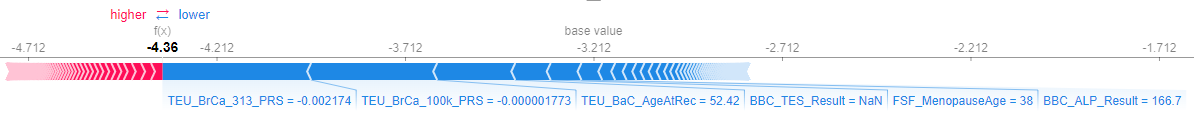

(a)

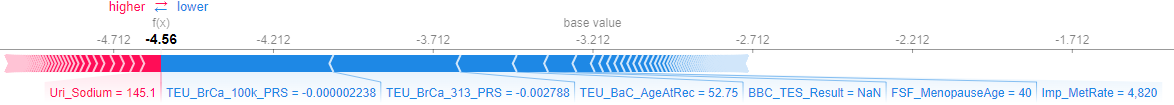

(b)

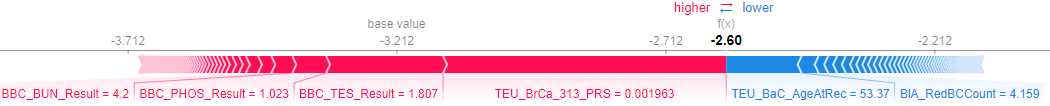

(c)

Supplementary Figure 9. Force plots explaining how much features influence the model prediction $f\left( \boldsymbol{x} \right)$ for three representative individuals with (a) ${SHAP}_{age}=-0.21$(b) ${SHAP}_{age}=-0.175$ (c) ${SHAP}_{age}=-0.14$. In each subplot, “base value” represents the average of all model predictions of the training data, hence the same for all samples. $f\left( \boldsymbol{x} \right)$ indicates model prediction (i.e. log (odds of developing breast cancer)). Features that contributed to the model prediction to a certain extent (i.e. SHAP value > 5% $\times\sum_{features} |SHAP values|$) were shown on the horizontal axis along with their raw values in the dataset. Red arrows indicates features which drive the odds of developing breast cancer higher, while blue arrows indicates the opposite. The length of the arrow reflects the amount of attribution of features (i.e. features with higher SHAP values have longer arrows). TEU_BrCa_313_PRS: PRS_313_. TEU_BrCa_100k_PRS: PRS_120k_. TEU_BaC_AgeAtRec: Age at recruitment, years. BBC_TES_Result: Testosterone. FSF_MenopauseAge: Age at menopause. BBC_ALP_Result: Alkaline phosphatase. Uri_Sodium: Sodium in urine. Imp_MetRate: Basal metabolic rate. BBC_BUN_Result: Plasma urea. BBC_PHOS_Result: Plasma phosphate. BlA_RedCCount: Red blood cell count.

The most obvious difference among three subplots in Supplementary Figure 9 is the drastic increase in model predictions from-4.36 to -2.60 at age 53. Among the features displayed in its subplots, the values of both PRS have the most distinctive change before and after the “jump” at age 53: PRS values increased from -0.002 to 0.002, which caused the swap in direction of the effect. Before the jump, the blue arrows of the PRS indicate protective effect on breast cancer, whereas the effect becomes opposite after the jump. Since both PRS have much higher importance (i.e. longer arrow) than others, we suspect PRS are responsible for the changes in model predictions before and after age of 53 years.

To confirm the increase in PRS across age 53, we plotted the histogram of PRS within this region by whether age is below 53 years (Supplementary Figure 10).

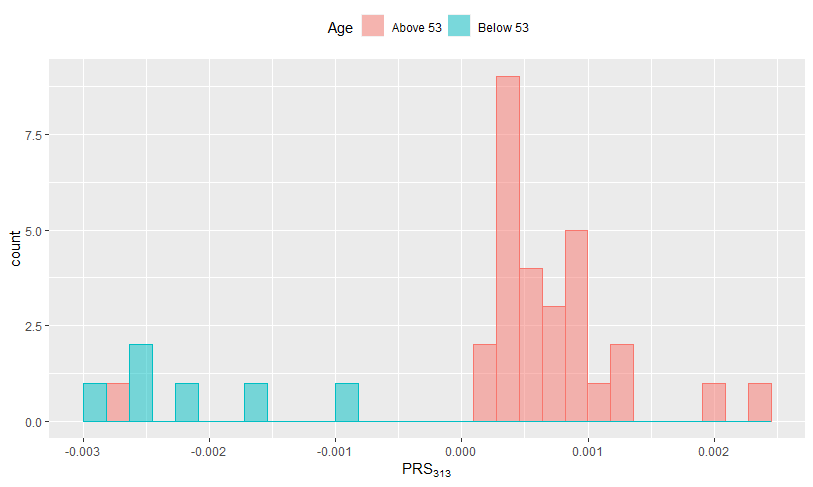

(a)

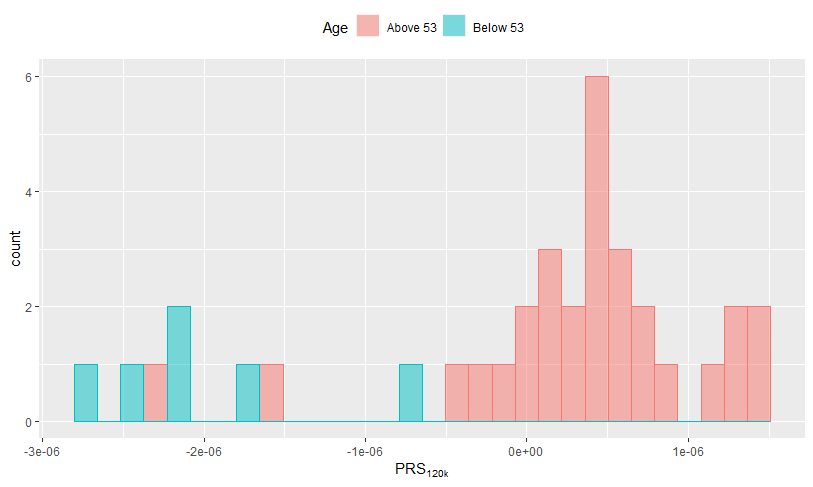

(b)

Supplementary Figure 10. Histogram of two breast cancer PRS by whether age below 53 among individuals with ${SHAP}_{age} \in(-0.21,-0.14)$ (a) PRS_313_ (b) PRS_120k_.

Supplementary Figure 10 confirms the increasing shift in PRS across age 53. As a comparison, we randomly picked another local region around age of 65 years of SHAP dependence plot, ${SHAP}_{age} \in(0.0814, 0.0815)$; its corresponding histogram (Supplementary Figure 11) suggested no shift in the distribution of PRS across age 65.

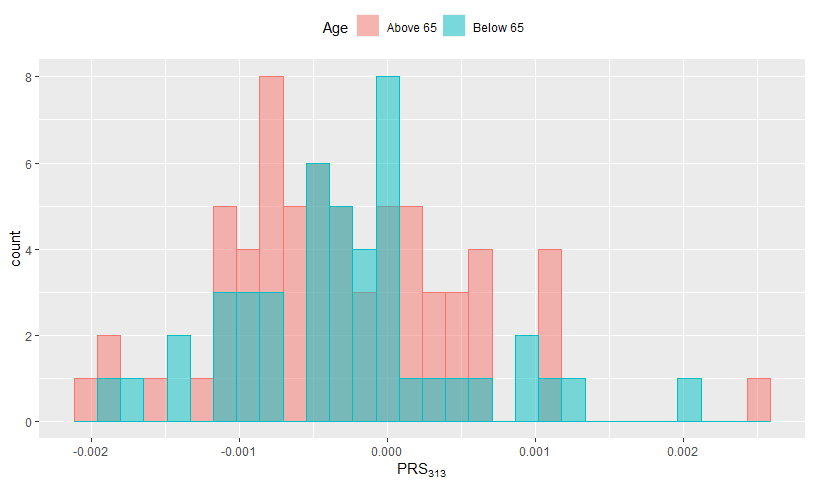

Supplementary Figure 11. Histogram of PRS_313_ by whether age below 56 among individuals with ${SHAP}_{age} \in(0.0814, 0.0815)$.

As a sensitivity analysis, we removed these 35 individuals and re-trained our XGBoost model; the “jump” persists.

In conclusion, our investigation thus far suggests that the “jump” in SHAP dependence plot is a reflection of the local pattern in our dataset where a subgroup of people had an increasing shift in PRS values at age 53. Since PRS are important features and increase in PRS is associated with higher risk of developing breast cancer, this provides the explanation of the jump in ${SHAP}_{age}$ at age 53. We emphasise that this local pattern should not be interpreted as a global behaviour such that the risk of developing breast cancer will suddenly increase after 53 years old.
